# Genetic subtypes of prediabetes, healthy lifestyle, and risk of type 2 diabetes: Prospective cohort study

**DOI:** 10.1101/2022.12.27.22283972

**Authors:** Yang Li, Guo-Chong Chen, Jee-Young Moon, Rhonda Arthur, Daniela Sotres-Alvarez, Martha L. Daviglus, Amber Pirzada, Josiemer Mattei, Krista M. Perreira, Jerome I. Rotter, Kent D. Taylor, Yii-Der Ida Chen, Sylvia Wassertheil-Smoller, Tao Wang, Thomas E. Rohan, Joel D. Kaufman, Robert Kaplan, Qibin Qi

**Affiliations:** Department of Epidemiology and Population Health, Albert Einstein College of Medicine, Bronx, NY 10461, USA; Department of Nutrition and Food Hygiene, School of Public Health, Soochow University, Suzhou, China; Department of Biostatistics, Gillings School of Global Public Health, University of North Carolina at Chapel Hill, Chapel Hill, NC 27599, USA; Institute for Minority Health Research, University of Illinois at Chicago, Chicago, IL 60612, USA; Department of Nutrition, Harvard T.H. Chan School of Public Health, Boston, MA 02115, USA; Department of Social Medicine, School of Medicine, University of North Carolina at Chapel Hill, Chapel Hill, NC 27599, USA; The Institute for Translational Genomics and Population Sciences, Department of Pediatrics, The Lundquist Institute for Biomedical Innovation at Harbor-UCLA Medical Center, Torrance, CA 90502, USA; Environmental & Occupational Health Sciences, Medicine, and Epidemiology, University of Washington, Seattle, WA 98105, USA

**Keywords:** prediabetes, type 2 diabetes, genetic subtyping, health lifestyle

## Abstract

**Objectives:** To cluster participants with prediabetes with five type 2 diabetes (T2D)-related partitioned polygenetic risk scores (pPRSs) and examine the risk of incident diabetes and the benefit of adherence to healthy lifestyle across clusters.

**Design:** Prospective cohort study

**Setting:** Hispanic Community Health Study/Study of Latinos (HCHS/SOL), US; UK Biobank (UKBB), UK.

**Participants:** 7,227 US Hispanic/Latinos without diabetes from HCHS/SOL, including 3,677 participants with prediabetes. 400,149 non-Hispanic whites without diabetes from UKBB, including 16,284 participants with prediabetes.

**Main outcome measures:** Prediabetes was defined by fasting plasma glucose (fasting glucose) between 100-125 mg/dL, 2-hour oral glucose tolerance test (OGTT 2h glucose) between 140-199 mg/dL, or hemoglobin A1c (HbA1c) between 5.7% and 6.5%. Diabetes was defined by fasting glucose levels ≥126 mg/dL, 2h glucose after OGTT ≥200 mg/dL, HbA1c ≥6.5%, current use of anti-diabetic medications, or medical record. Five pPRSs representing various pathways related to T2D were calculated based on 94 T2D-related genetic variants. Health lifestyle score was assessed with five modifiable risk factors, including body mass index (BMI), smoking, alcohol drinking, physical activity, and diet for T2D.

**Results:** Using K-means consensus clustering on five pRPSs, six clusters of individuals with prediabetes were identified in HCHS/SOL, with each cluster presenting disparate patterns of pPRSs and different patterns of metabolic traits. Except cluster 3 which was not detected, the other five clusters were conformed in participants with prediabetes in UKBB, with each cluster showing the similar patterns of pPRSs to their corresponding cluster in HCHS/SOL. At baseline, proportion of impaired glucose tolerance (IGT)/impaired fasting glucose (IFG) and glycemic traits in HCHS/SOL (fasting glucose, OGTT 2h glucose, and HbA1c) were not significantly different across six clusters (P=0.13, P=0.62, P=0.35, P=0.96, respectively). In UKBB, random glucose and HbA1c at baseline did not show significant difference across five clusters (P=0.43, P=0.71, respectively). Although baseline glycemic traits were similar across clusters, cluster 6, which featured a very low proinsulin score, exhibited elevated risk of incident T2D in both cohorts (risk ratio [RR]=1.39, 95% confidence interval [95% CI]=[1.10, 1.76] vs. cluster 1 in HCHS/SOL; hazard ratio [HR]=1.29, 95% CI=[1.00, 1.69] vs. cluster 1 in UKBB; Combined RR/HR=1.34 [1.13, 1.60]). To explain the elevated risk of incident T2D in cluster 6, interactions between proinsulin score and other three pPRSs (Beta-cell score, Lipodystrophy-like score, Liver-lipid score) and sum score were detected (P for interaction=0.001, 0.04, 0.02 and 0.002, respectively). Cluster 5 showed an increased risk of incident T2D in UKBB (HR=1.35 [1.05, 1.75] vs. cluster 1) and in the combined analysis with HCHS/SOL (RR/HR=1.29 [1.08, 1.53]), although its risk of T2D was not significantly different from cluster 1 in HCHS/SOL (RR=1.23 [0.96, 1.57]). Inverse associations between the lifestyle score and risk of T2D were observed across different clusters, with a suggestively stronger association in Cluster 5 compared to Cluster 1, in both cohorts. Cluster 5 showed reduced risk of incident diabetes caused by healthy lifestyle score (RR=0.65 [0.47, 0.89], HR=0.71 [0.62, 0.81], respectively. Combined RR/HR=0.70 [0.62, 0.79]). Among individuals with a healthy lifestyle, those in Cluster 5 had a similar risk of T2D compared to those in Cluster 1 (combined RR/HR=1.03 [0.91-1.18], P>0.05).

**Conclusions:** This study identified genetic subtypes of prediabetes which differed in risk of progression to T2D, with two subtypes showing relatively high risk of T2D over time. Favorable relationship between healthy lifestyle and risk of T2D was observed, regardless of their genetic subtypes. Participants in one subtype with higher risk of T2D may realize extra benefits in terms of risk reduction from a healthy lifestyle.

## Introduction

Prediabetes is a condition of impaired glucose metabolism often preceding the ascertainment of overt diabetes.^1^ The term “prediabetes” can refer to a clinical state defined alternatively upon an observed fasting glucose between 5.6-7.0 mmol/L (impaired fasting glucose [IFG]), a glucose level measured 2h after oral glucose tolerance test (OGTT) between 7.8-11.1 mmol/L (impaired glucose tolerance [IGT]), or an HbA1c level between 5.6-6.5%.^2^ Although all these subtypes carry the same designation of “prediabetes”, prediabetes is a heterogenous metabolic state that varies in many aspects of its pathogenesis.^3^ In turn, subtypes of prediabetes may differ in their associated risks of future development of type 2 diabetes (T2D) and its complications, such as cardiovascular diseases (CVD) and kidney disease.^3^ In addition to the standard approach to prediabetes classification based upon glucose and HbA1c categories, additional metabolic features have been proposed to improve the classification of prediabetes. Wagner et al. used phenotypic variables related to glucose tolerance, insulin sensitivity, insulin secretion, lipids, adiposity, and a T2D polygenic risk score to identify six clusters of subphenotypes in persons without diabetes.^4^ Individuals in one cluster characterized by insulin resistance and fatty liver had the highest risk of T2D, and individuals in another cluster characterized by high visceral fat had the most increased risk of kidney disease and all-cause mortality.^4^ These previous studies demonstrated the pathophysiological heterogeneity among individuals before diagnosis of T2D, thus suggesting that subtyping of prediabetes based on multiple features could help to improve stratification of disease risk.^3^

Previous efforts to subtype prediabetes and T2D have mainly focused on phenotypic variables,^3-6^ while many T2D-associated genetic variants have been identified through GWAS^7-10^, with various pathophysiological pathways.^8^ Based on associations between 94 T2D-associated variants and 47 T2D-related traits, Udler et al. created five pathway-based partitioned polygenetic risk scores (pPRSs) representing two mechanisms of beta-cell dysfunction (i.e., beta-cell dysfunction score and low proinsulin score) and three mechanisms of insulin resistance (i.e., obesity score, lipodystrophy-like score, and disrupted liver lipid metabolism score) in relation to T2D.^11^ A recent GWAS of five subtypes of diabetes, defined by six clinical variables,^5^ also suggested that diabetes subtypes may have partially distinct genetic drivers indicating etiological differences.^12^ However, genetic information reflecting different pathophysiological pathways has not been well-utilized in subtyping individuals at elevated risk of T2D.^4^ Compared to phenotypic variables, germline genetic variants are more likely to point to disease causation and do not change with disease progression or treatment.^11^ Thus, subtyping of prediabetes based on genetic variants with different pathophysiological pathways will not only help better understand pathophysiological heterogeneity but also provide useful information in early prediction and prevention before onset of clinically detectable metabolic changes and diagnosis of T2D.

In this study, we aimed to subtype prediabetes based on five previously reported pPRSs reflecting different genetic pathways of T2D in a prospective cohort of diverse Hispanic/Latino individuals with higher burden of diabetes.^13^ We then compared risk of incident T2D among individuals in different clusters of prediabetes during an average six-year follow-up. In addition, we also examined associations of healthy lifestyle and risk of T2D across different clusters of prediabetes. We replicated the analysis in a large, independent, prospective cohort of European non-Hispanic white adults from the UK Biobank (UKBB).^14^

## Methods

### Study population

The Hispanic Community Health Study/Study of Latinos (HCHS/SOL) is a population-based cohort that recruited 16,415 adults living in four US metropolitan areas who were self-identified as Hispanic or Latino and were aged 18-74 years at baseline.^15 16^ A comprehensive battery of interviews and a clinical assessment including fasting and post-OGTT blood draw with central analysis of laboratory analytes were conducted at in-person clinic visits during 2008-2011 (baseline) and 2014-2017 (Visit 2). The current analysis included 7,227 participants who were free of diabetes at baseline, had genetic data, and completed the six-year follow-up (Visit 2) examination. The study was approved by the Institutional Review Boards at all participating institutions. All participants gave written informed consent.

UKBB is a large prospective observational study that recruited approximately 500,000 men and women of various ethnicities aged 37-73 years from across 22 centers located throughout England, Wales, and Scotland between 2006 and 2010.^14^ Participants underwent various measurements including a standardized baseline laboratory assessment and provided a wide range of information on health and diseases at recruitment. In the current analysis, 400,149 non-Hispanic white participants without diabetes at baseline and with genetic data were included, with a median 10.1 years of follow-up. UKBB received ethical approval from the research ethics committee (REC reference for UKBB 11/NW/0382), and participants provided written informed consent. The current study was conducted under application number 56483 of UKBB resource. An expanded description of the methods is available in the Supplementary Appendix.

### Ascertainment of prediabetes and diabetes

In HCHS/SOL, prediabetes was defined as fasting glucose between 100-125 mg/dL, 2h glucose after OGTT between 140-199 mg/dL, or HbA1c between 5.7% and 6.5%, and no self-reported physician-diagnosed diabetes or medically treated diabetes.^2^ Diabetes was defined as fasting glucose levels ≥126 mg/dL, 2h glucose after OGTT ≥200 mg/dL, HbA1c ≥6.5%, or current use of anti-diabetic medications.^2^ Participants free of diabetes at baseline (Visit 1) who were identified as having diabetes at Visit 2 were deemed to be incident cases of T2D.

In UKBB, diabetes was defined through multiple procedures and sources of the diagnosis including primary care, hospital admissions, and self-reported, or participants with HbA1c ≥ 6.5% according to UKBB algorithms to determine diabetes status. ^2 14 17^ Individuals with prediabetes were defined as those with HbA1c between 5.7% and 6.5%.^2^ Participants free of diabetes at baseline who were identified as having diabetes during follow-up were deemed to be incident cases of T2D.

### Measurements of metabolic traits

In HCHS/SOL, BMI was calculated as weight in kilograms divided by height in meters squared. Waist-to-hip ratio (WHR) is the ratio of the circumference of the waist to that of the hips. Centralized laboratory tests were performed to determine plasma fasting glucose, 2h glucose after OGTT, fasting insulin, and hemoglobin A1c (HbA1c), and serum lipids including triglycerides, and total, low-density lipoprotein (LDL), and high-density lipoprotein (HDL) cholesterol. Homeostatic model assessment of insulin resistance (HOMA-IR) and beta-cell function (HOMA-B) were derived using equations based on fasting glucose and insulin.^18^ In UKBB, metabolic traits included BMI, WHR, random glucose, HbA1c, triglycerides, and total-, LDL- and HDL-cholesterol.^19^

### Genotyping, pPRSs calculation and clustering

In HCHS/SOL, genome-wide genotyping was performed in 12,633 participants using a customized Illumina array (15041502 B3; Illumina Omni 2.5M array plus ∼150K custom SNPs), and imputation was carried out based on the 1000 Genomes Project phase 3 reference panel.^20^ In UKBB, genome-wide genotyping was performed in ∼500,000 participants using Affymetrix UK BiLEVE Axiom Array or Affymetrix UKBB Axiom Array, and imputation was carried out based on the HRC reference (Version 3 imputed data).^21^ Five pPRSs, including beta-cell score, proinsulin score, obesity score, lipodystrophy-like score, and liver-lipid score, were calculated based on 94 T2D-assocaited genetic variants using a previously reported method.^11^ Briefly, 94 variants were partitioned into five SNP sets in Bayesian nonnegative matrix factorization (bNMF) with the partitioning process being supervised by 47 T2D-related traits. Within each SNP set, a polygenetic risk score was assessed by computing the sum of risk alleles of SNPs in the set for each individual, weighted by their effect sizes estimated by previous study.^11^ Thus, five PRSs were calculated for each individual as input features for the following clustering analysis on participants with prediabetes.

To cluster participants with prediabetes, K-means consensus clustering was performed on 3,677 participants with prediabetes in HCHS/SOL and on 16, 284 participants with prediabetes in UKBB, using the ConsensusClusterPlus R package, respectively.^22^ A subset of participants were selected, and Pearson’s correlation coefficient among them was calculated using five pPRSs. Then, K-means clustering was performed based on the correlation matrix, with each participant being assigned into one cluster. The whole process was repeated 1,000 times. The cluster memberships of the single clustering result were combined to a consensus matrix, determining the number of times each pair of samples was clustered together over all clusterings. The following settings were used for clustering: number of repetitions = 1,000 bootstraps; pItem = 0.8 (resampling 80% of any sample); pFeature = 1.0 (using all pPRSs); and the number of whole clusters was set from two to 12. The best number of clusters was determined by the largest average Silhouette coefficients.

### Lifestyle score calculation

Adherence to a healthy lifestyle was measured by a lifestyle score based on five well-established modifiable factors including BMI, smoking, alcohol drinking, physical activity, and diet components relevant to risk of T2D.^23^ Each factor was scored individually, and then the overall lifestyle score was calculated by summing up five scores (**Supplementary Table 1 and 2**). In HCHS/SOL, both smoking and drinking status were measured by self-report and categorized into three categories: current, former, and never. Participants who reported current alcohol drinking were asked to provide the number of drinks consumed in a week. Physical activity was measured using the Global Physical Activity Questionnaire.^24^ Dietary intake was estimated using the National Cancer Institute methodology based on two 24-h dietary recalls and a food propensity questionnaire.^25^ Dietary quality was assessed using the Alternative Healthy Eating Index (AHEI)-2010.^26^

In UKBB, similar approaches were used to measure lifestyle factors and calculate the lifestyle score (**Supplementary Table 2**). Since the AHEI-2010 was not available in UKBB, we assessed overall dietary quality based on five major food groups associated with T2D according to previous studies.^27-29^ For both studies, participants within the 2^nd^ and 3^rd^ tertiles of the lifestyle score were defined as being adherent to a healthy lifestyle.

## Statistical Analysis

After calculating pPRSs, linear regression was used to examine associations of five pPRSs with baseline metabolic traits after adjustment for covariates in both cohorts. To examine the association between five pPRSs and risk of T2D (per SD increment) among participants without diabetes at baseline, Robust Poisson regression adjusting for covariates was performed in HCHS/SOL, while Cox proportional-hazards regression adjusting for covariates was used in UKBB.

After obtaining clusters of prediabetes, Analysis of Variance (ANOVA) and Chi-squared tests were applied to test differences in five pPRSs and characteristics including continuous variables (e.g., baseline glycemic traits and changes in glycemic traits) and categorical variables (e.g., glycemic status [impaired glucose tolerance (IGT), only impaired fasting glucose (IFG), or both] and Hispanic/Latino background) across clusters of prediabetes in both cohorts. To compare the relative risks of T2D across clusters of prediabetes (with Cluster 1 defined as reference), Robust Poisson regression and Cox proportional-hazards regression adjusting for covariates were performed in HCHS/SOL and UKBB, respectively.

To detect the interaction between proinsulin score and the other four pPRSs and a summed score (summing up these four pPRSs), Robust Poisson regression and Cox proportional-hazards regression adjusting for covariates were performed after participants without diabetes were stratified by the median of the proinsulin score (interactions were test by including the respective interaction terms in the models) in HCHS/SOL and UKBB, respectively.

The associations between adherence to a healthy lifestyle and risk of T2D in all participants with prediabetes, as well as across clusters of prediabetes, were estimated using Robust Poisson regression and Cox proportional-hazards regression adjusting for covariates in HCHS/SOL and UKBB, respectively. The relative risks of T2D comparing Cluster 5 to Cluster 1 while stratifying according to the lifestyle score (with Cluster 1 with unhealthy lifestyle defined as reference) were also assessed in both cohorts using Robust Poisson regression and Cox proportional-hazards regression adjusting for covariates, respectively.

For all the analyses above, RRs and 95% confidence intervals were reported when using Robust Poisson regression in HCHS/SOL, and HRs and 95% confidence intervals were reported when using Cox proportional-hazards regression in UKBB. Results from HCHS/SOL and UKBB were pooled by inverse-variance weighted, fixed-effects meta-analyses.

In the combined analysis, difference in the effect sizes of healthy lifestyle on risk of T2D between clusters was tested by Cochran’s Q test for heterogeneity. We considered 2-sided *P* values <0.05 as statistically significant and used False Discovery Rate (FDR) adjusted P value for multiple tests. All analyses were performed using R 4.1.2.

## Results

### Baseline characteristics of participants

The baseline characteristics of the participants from HCHS/SOL and UKBB are shown in **Supplementary Table 3 and Supplementary Table 4**. Differences in the characteristics between participants with normal glucose and those with prediabetes were generally similar for the two studies. Compared to those with normal glucose, participants with prediabetes were older, more likely to be males and current smokers and had lower education levels. Participants with prediabetes had higher levels of obesity measures, glycemic traits, and less favorable lipid profiles (i.e., higher triglycerides, lower HDL-cholesterol) compared to those with normal glucose.

### Genetic risk scores, metabolic traits, and risk of T2D

We first calculated five pPRSs, including the beta-cell score, proinsulin score, obesity score, lipodystrophy-like score and liver-lipid score, among 7,227 participants without diabetes at baseline in HCHS/SOL and 400,149 participants without diabetes at baseline in UKBB (**Supplementary Figure 1**). Relatively weak correlations were observed among these five pPRSs both in HCHS/SOL (r=0.07 to 0.26) and in UKBB (r=0.10 to 0.28) (**Supplementary Figure 2)**. In HCHS/SOL, beta-cell score and low proinsulin score were inversely associated with beta-cell function measured by HOMA-B, while obesity score, lipodystrophy-like score and liver-lipid score were positively associated with insulin resistance measures (i.e., HOMA-IR and fasting insulin) (**Supplementary Figure 3A)**. All five scores were positively correlated with fasting glucose, 2h glucose after OGTT and HbA1c. Previously reported associations of these pPRSs with other corresponding phenotypic traits^17^ were also confirmed in HCHS/SOL (e.g., higher obesity score was correlated with higher BMI). Similar correlations of these pPRSs with glycemic traits (e.g., random glucose, HbA1c) and other metabolic traits were observed in UKBB (**Supplementary Figure 3B)**, although HOMA-B and HOMA-IR were not measured. We then examined associations of these pPRSs with incident T2D. Among 7,227 participants without diabetes at baseline in HCHS/SOL, 888 incident T2D cases were identified after an average of 6-year follow-up. Among 400,149 participants without diabetes at baseline in UKBB, 10,806 incident T2D cases were identified during a median 10.1 years of follow-up. All five scores were positively associated with the risk of T2D in both studies (all *P*<0.05) (**Supplementary Figure 3 C and D**).

### Genetic subtypes of prediabetes and baseline metabolic profiles

Based on the five pPRSs, we used K-means-based consensus clustering to classify genetic subtypes among 3,677 participants with prediabetes at baseline in HCHS/SOL. Six clusters of prediabetes were identified, which was determined according to the highest average Silhouette coefficients (**Supplementary Figure 4**). The numbers of individuals within each cluster from Cluster 1 to Cluster 6 were 488 (13.2%), 569 (15.5%), 547 (14.8%), 742 (20.0%), 719 (19.5%) and 612 (16.6%), respectively, and each cluster displayed distinctive pPRS patterns (**Figure 1A**). For example, Cluster 1 had a very low (far below average) beta-cell score, high (above average) levels of obesity score and liver-lipid score, and average levels of low proinsulin score and lipodystrophy-like score. Metabolic trait patterns were generally consistent with the pPRS patterns in each cluster (**Figure 1B**). For example, Cluster 1 had high levels of HOMA-B (corresponding to low beta-cell score), BMI and HOMA-IR (corresponding to high levels of obesity score and liver lipid score). Differences in the pPRSs and metabolic traits between any two clusters of prediabetes are shown in **Supplementary Figure 5**. Main features of the pPRSs and metabolic traits for these six clusters are summarized in **Supplementary Table 5**. We then used the same consensus clustering approach and identified five genetic subtypes among 16,284 participants with prediabetes at baseline in UKBB **(Supplementary Figure 4)**. These clusters were highly consistent with those identified in HCHS/SOL, except for Cluster 3 which was not detected in UKBB (**Figure 1C)**.

**Figure 1.**
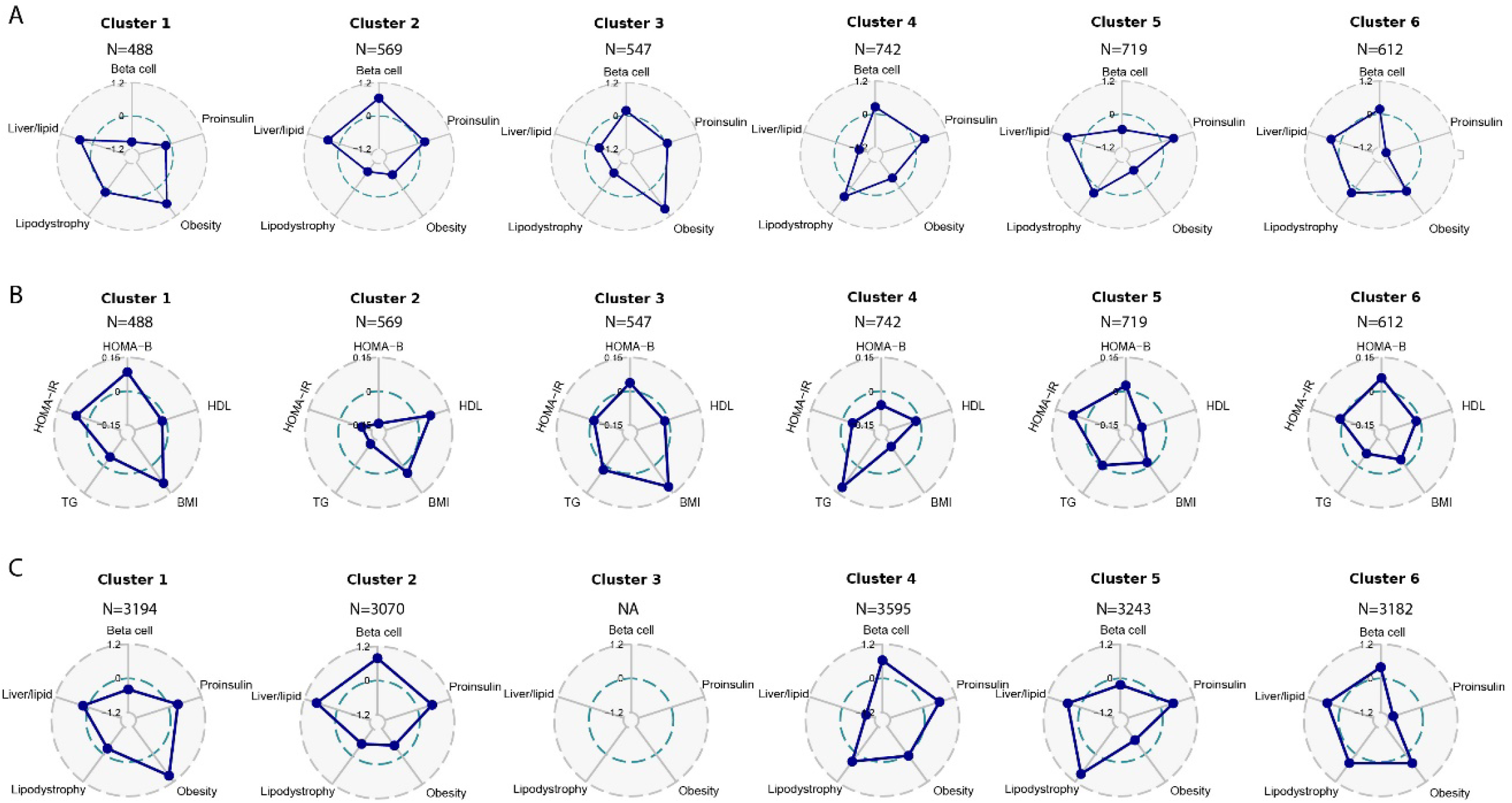
Patterns of genetic scores and metabolic traits pattern across clusters of prediabetes. **(A)** Patterns of five pPRSs across six clusters of individuals with prediabetes in HCHS/SOL. **(B)** Patterns of five T2D-related metabolic traits across six clusters of individuals with prediabetes in HCHS/SOLHCHS/SOL. **(C)** Patterns of five pPRSs across five clusters of individuals with prediabetes in UKBB (Cluster 3 was not detected in UKBB). Blue dots on five axes in Radar plot show the median values of standardized pPRSs or standardized metabolic traits.

Although these clusters had distinctive metabolic trait pattern corresponding to their various pPRS patterns, after examining the standard clinical markers of glycemic status, we concluded that none of the clusters showed an overall better or worse metabolic pattern at baseline than others (**Supplementary Table 6 and Supplementary Table 7**). In particular, proportions of individuals with IFG, IGT or both were similar (*P*=0.13), and levels of fasting glucose (*P*=0.62), 2h glucose after OGTT (*P*=0.35) and HbA1c (*P*=0.98) were similar across 6 clusters in HCHS/SOL (**Figure 2A-D**). In UKBB, levels of random glucose (*P*=0.43) and HbA1c (*P*=0.71) were also similar across 5 clusters (**Figure 2E-F)**. There were no differences in demographic (e.g., age, sex), socioeconomic (education, family income), and behavioral (e.g., smoking, drinking) factors across clusters in either study (**Supplementary Table 6 and Supplementary Table 7**), except for Hispanic/Latino background in HCHS/SOL. However, all six clusters were identified in each of the six Hispanic/Latino groups, and no group was dominated by one or two clusters (**Supplementary Figure 6)**.

**Figure 2.**
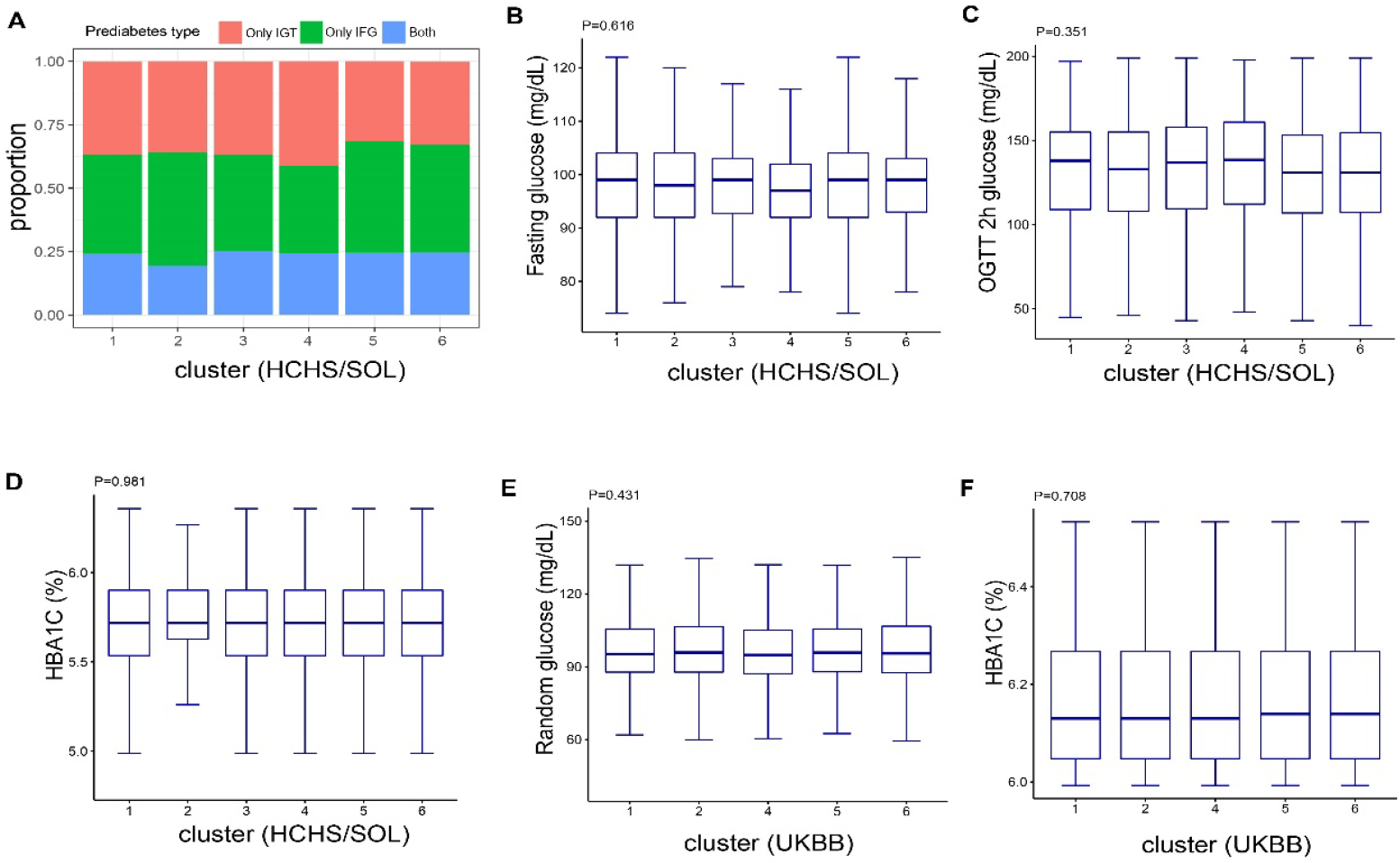
Comparison of baseline metabolic traits and glycemic status across clusters of prediabetes. **(A)** Proportion of individuals with only impaired glucose tolerance (IGT), only impaired fasting glucose (IFG), or both across six clusters in HCHS/SOL. **(B-D)** Boxplots of fasting glucose, 2h glucose after OGTT and HbA1C at baseline across six clusters in HCHS/SOL. **(E-F)** Boxplots of random glucose and HbA1C at baseline across five clusters in UKBB.

### Genetic subtypes of prediabetes and incident T2D

We then compared risk of incident T2D among these clusters of individuals with prediabetes at baseline. In HCHS/SOL, individuals in Cluster 6 characterized by a very low proinsulin score had the highest risk of T2D (RR=1.39 [1.10, 1.76] compared to Cluster 1; *P*=0.006), followed by those in Cluster 5 characterized by high levels of liver-lipid score, lipodystrophy-like score and proinsulin score, and low levels of beta-cell score and obesity score (RR= 1.23 [0.96, 1.57] compared to Cluster 1, *P*=0.096) (**Figure 3A**). In UKBB, individuals in Cluster 5 and Cluster 6 also showed the highest risk of T2D compared to Cluster 1 (HR= 1.35 [1.05, 1.75] and 1.29 [1.00, 1.67], *P*= 0.021 and 0.049, respectively). In the combined analysis of two cohorts, individuals in Cluster 5 and Cluster 6 had 29% [8%, 53%] and 34% [13%, 60%] increased risk of T2D compared to those in Cluster 1 (*P*=0.005 and <0.001, respectively).

We also examined changes in glycemic traits over six years across these six clusters in HCHS/SOL and found that changes in fasting glucose (**Figure 3B**; overall *P*=0.02), 2h glucose after OGTT (**Figure 3C;** overall *P*=0.003) and HOMA-IR (**Figure 3D**; overall *P*=0.03) differed across clusters, despite baseline levels of these traits being similar across clusters. Compared to those in Cluster 1, individuals in Cluster 5 and Cluster 6 had greater increase in fasting glucose and 2h glucose after OGTT over 6 years (all *P*<0.05) (**Figure 3B-C**).

We considered it unexpected that individuals with prediabetes in Cluster 6, which stood out for having a very low proinsulin score, with only slightly above average levels of other four scores, showed the highest risk of T2D (**Figure 1A and Figure 3A**), since lower proinsulin score was associated with lower risk of T2D and favorable glycemic traits, including higher HOMA-B and lower levels of fasting glucose, 2h glucose after OGTT and HbA1c. (**Supplementary Figure 3**). Revisiting the associations of the proinsulin score and other genetic scores with risk of T2D, we found evidence of interaction between the proinsulin score and other genetic scores on risk of T2D (**Figure 4**). In HCHS/SOL, the associations of Beta-cell score and Liver/Lipid score with risk of T2D were stronger among individuals with a low proinsulin score, than among those with a high proinsulin score (P for interaction was 0.04 and 0.06 respectively). The combined genetic effect of other four scores (calculated as a summed score without the proinsulin score) was stronger among individuals with a low proinsulin score (RR=1.31, [95% CI 1.20, 1.42], P<0.001) compared to those with a high proinsulin score (RR=1.09 [95% CI 0.99, 1.21], P=0.09) (P for interaction =0.01). These potential interactions were replicated in UKBB (all *P* for interaction <0.05), with an additional interaction between lipodystrophy-like score and proinsulin score identified (P for interaction=0.01).

**Figure 3.**
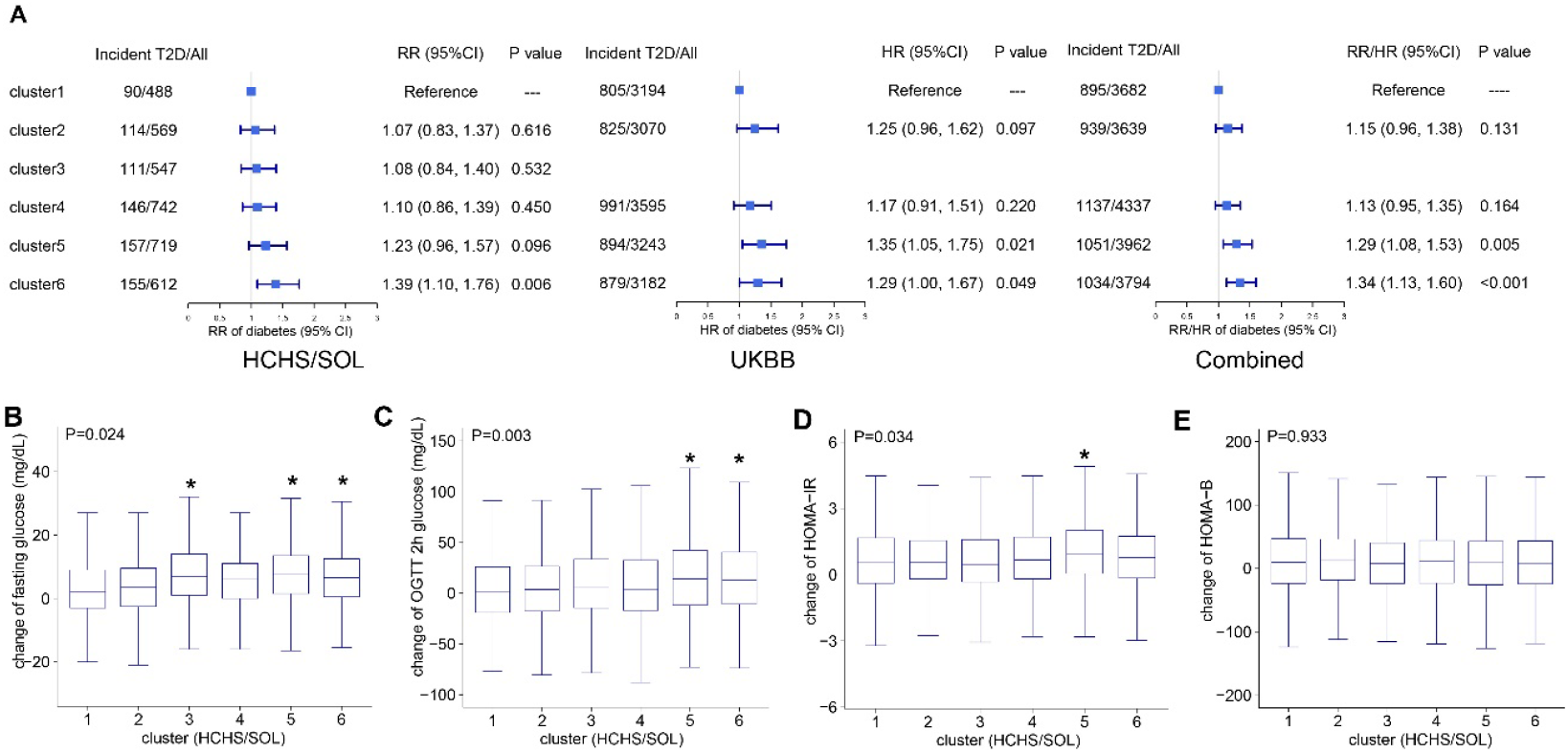
Clusters of prediabetes, incident T2D and changes in glycemic traits during follow-up. **(A)** Clusters of prediabetes and risk of T2D in HCHS/SOL, UKBB, and the combined studies. In HCHS/SOL, data are risk ratios (RRs) and 95% confident intervals (CIs) estimated by Poisson regression after adjustment for age, gender, U.S.-born status, Hispanic/Latino background, education, annual income, AHEI-2020, smoking, drinking, physical activity, and eigenvectors derived from GWAS. In UKBB, data are Hazard ratios (HRs) and 95% CIs estimated by Cox proportional-hazards regression after adjustment for age, gender, education, Townsend deprivation score, diet score, smoking, drinking, physical activity, and eigenvectors derived from GWAS. In the combined analysis, results from HCHS/SOL and UKBB were combined using fixed-effect meta-analysis. **(B-E)** Boxplots of changes in fasting glucose, 2h glucose after OGTT, HOMA-IR and HOMA-B over 6 years across six clusters of individuals with prediabetes in HCHS/SOL.

**Figure 4.**
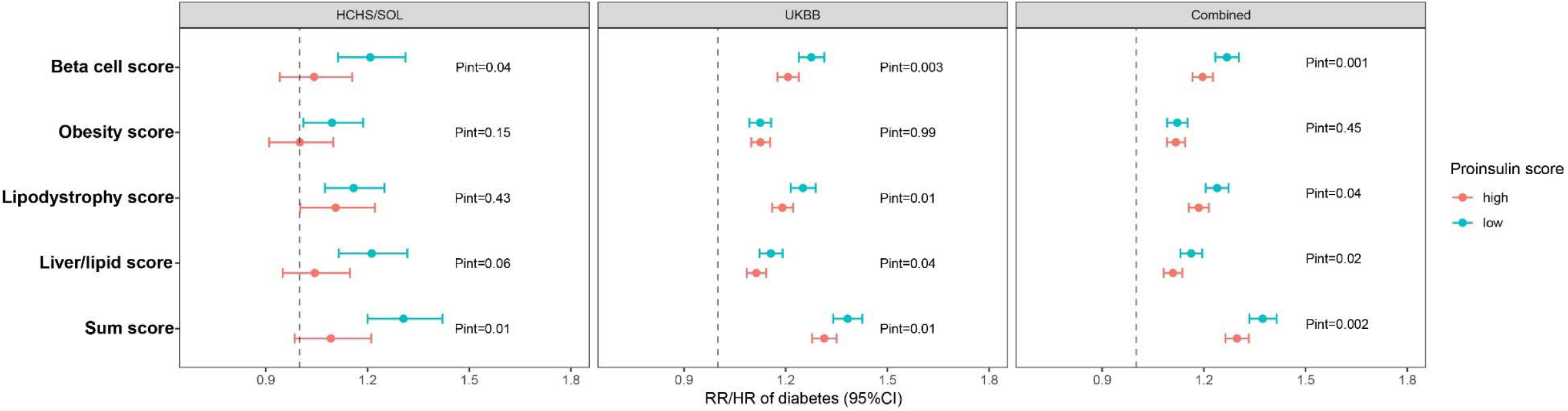
Associations of other genetic scores with incident T2D according to proinsulin score. In HCHS/SOL, data are risk ratios (RRs) and 95% confident intervals (CIs) for incident T2D per SD increment of the corresponding genetic scores, estimated by Poisson regression after adjustment for age, gender, U.S.-born status, Hispanic/Latino background, education, annual income and eigenvectors derived from GWAS. In UKBB, data are Hazard ratios (HRs) and 95% CIs for incident T2D per SD increment of the corresponding genetic scores, estimated by Cox proportional-hazards regression after adjustment for age, gender, education, Townsend deprivation score and eigenvectors derived from GWAS. In the combined analysis, results from HCHS/SOL and UKBB were combined using fixed-effect meta-analysis. High and low proinsulin score was defined by the median. The sum score was calculated by summing up Beta cell score, Obesity score, Lipodystrophy-like score, and Liver-lipid score.

### Healthy lifestyle, genetic subtypes of prediabetes and risk of T2D

We then examined the association between adherence to a healthy lifestyle, estimated by a lifestyle score based on BMI, alcohol intake, smoking, physical activity, and diet in HCHS/SOL (**Supplementary Table 1**) and in UKBB (**Supplementary Table 2**), and risk of T2D across different clusters of individuals with prediabetes (**Figure 5A**). In HCHS/SOL, a healthy lifestyle (i.e. 2^nd^ and 3^rd^ tertiles of the lifestyle score) was associated with a decreased risk of T2D in all individuals with prediabetes (RR=0.73 [0.63, 0.84], *P*<0.001), among those in Cluster 2 (RR=0.67, [0.47, 0.95], *P*=0.02) and those in Cluster 5 (RR=0.65, [0.47, 0.89], *P*=0.008), but not among those in other clusters. In UKBB, a healthy lifestyle was significantly associated with a decreased risk of T2D in all individuals with prediabetes (RR=0.80 [0.75, 0.85], *P*<0.001) as well as in each cluster separately, with the highest effect size observed in Cluster 5 (RR=0.71, [0.62, 0.81], *P*<0.001) and the lowest effect size observed in Cluster 1 (RR=0.86, [0.75, 1.00], *P*=0.048). In the combined analysis of two cohorts, the inverse association between healthy lifestyle and risk of T2D was observed across all five clusters, with a stronger association observed in Cluster 5 (RR=0.70, [0.62, 0.79], P<0.001) compared to Cluster 1 (RR=0.85, [0.74, 0.97], P=0.02). We did not found overall difference across all clusters using Cochran’s Q test for heterogeneity. However, a different effect size was detected between cluster 1 and cluster 5 (P=0.04 for difference in the effect size).

**Figure 5.**
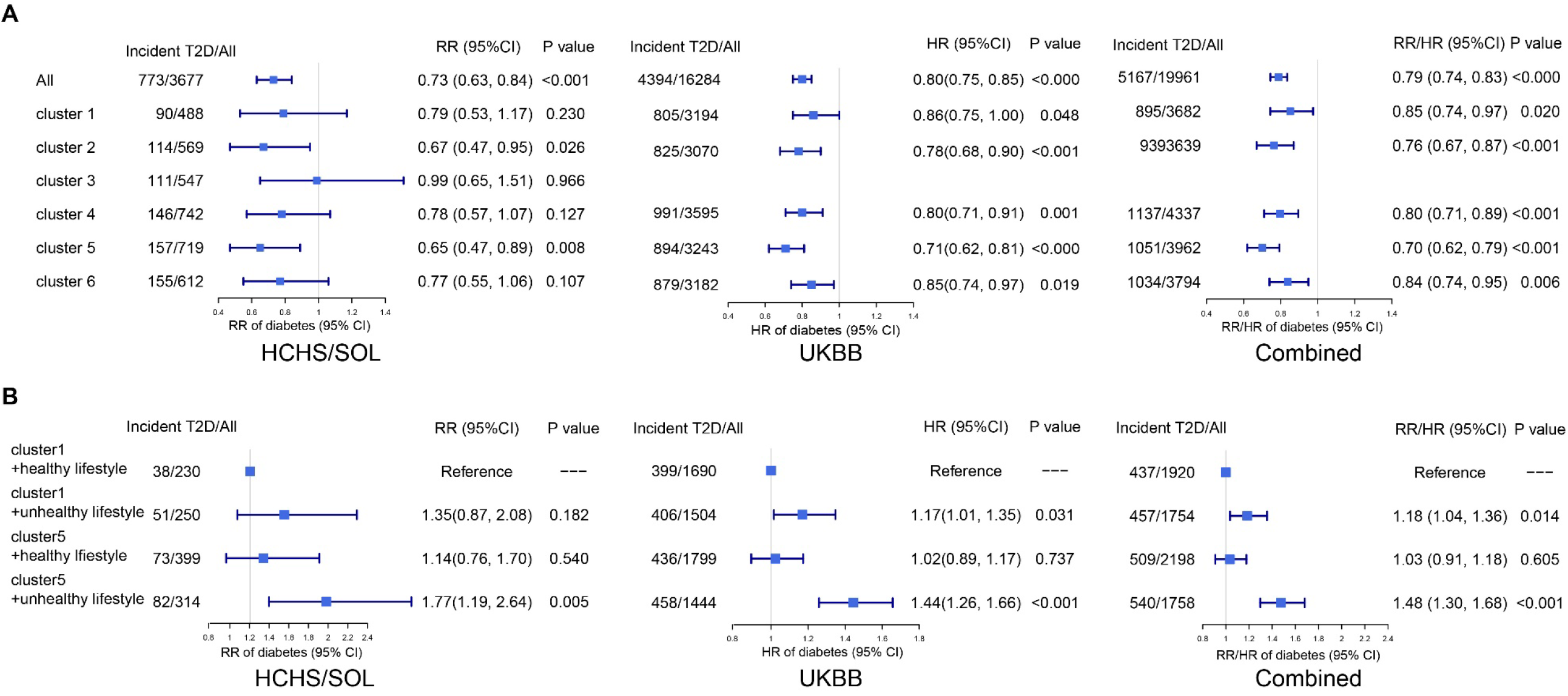
Association between healthy lifestyle and incident T2D across clusters of prediabetes. (A) Association between healthy lifestyle and incident T2D among all individuals with prediabetes and across clusters of prediabetes. In HCHS/SOL, data are risk ratios (RRs) and 95% confident intervals (CIs) for incident T2D by comparing healthy lifestyle (2nd and 3rd tertiles of the lifestyle score) with unhealthy lifestyle (the 1st tertile of the lifestyle score), estimated by Poisson regression after adjustment for age, gender, U.S.-born status, Hispanic/Latino background, education, annual income and eigenvectors derived from GWAS. In UKBB, data are Hazard ratios (HRs) and 95% CIs for incident T2D by comparing healthy lifestyle (2nd and 3rd tertiles of the lifestyle score) with unhealthy lifestyle (the 1st tertile of the lifestyle score), estimated by Cox proportional-hazards regression after adjustment for age, gender, education, Townsend deprivation score and eigenvectors derived from GWAS. In the combined analysis, results from HCHS/SOLand UKBB were combined using fixed-effect meta-analysis. (B) Risk of T2D in the Cluster 5 compared to the Cluster 1 according to lifestyle (healthy lifestyle: 2nd and 3rd tertiles of the lifestyle score; unhealthy lifestyle: 1st tertile of the lifestyle score). In HCHS/SOL, data are risk ratios (RRs) and 95% CIs for incident T2D, estimated by Poisson regression after adjustment for covariates mentioned above. In UKBB, data are Hazard ratios (HRs) and 95% CIs for incident T2D, estimated by Cox proportional regression after adjustment for covariates mentioned above. In the combined analysis, results from HCHS/SOL and UKBB were combined using fixed-effect meta-analysis.

To further illustrate potentially extra beneficial effects of health lifestyle on risk of T2D in Cluster 5 compared to Cluster 1, we compared risk of T2D between Cluster 1 and Cluster 5 according to the lifestyle score (**Figure 5B**). In HCHS/SOL, compared to those in Cluster 1 with a healthy lifestyle, individuals in Cluster 5 with an unhealthy lifestyle had a much higher risk of T2D (RR=1.77 [1.19, 2.64], *P*=0.005), while those in Cluster 5 with a healthy lifestyle did not show a significant higher risk of T2D (RR=1.14 [0.76, 1.70], *P*=0.54). Similar results were observed in UKBB. In the combined analysis of two cohorts, given a healthy lifestyle, individuals in Cluster 5 had a similar risk of T2D compared to those in Cluster 1(RR=1.03 [0.91, 1.18], *P*=0.60).

## Discussion Principle findings

Based on five genetic risk scores representing different pathophysiological pathways related to T2D, we identified six clusters of prediabetes, with distinctive patterns of genetic risk scores and corresponding metabolic traits in US Hispanic/Latinos from HCHS/SOLand confirmed five clusters in non-Hispanic white adults from UKBB. Different clusters of individuals with diabetes had similar levels of blood glucose and HbA1c at baseline but differed in risk of progression to T2D during follow-up.

### Comparison with other studies and mechanistic insights

Clusters of prediabetes identified by our approach exhibited distinctive patterns of genetic scores and corresponding patterns of diabetes-related metabolic traits which reflect beta-cell function, insulin resistance, obesity and lipid metabolism. However, interestingly, baseline glycemic traits (e.g., fasting and 2h post oral load glucose levels, HbA1c) and proportions of individuals with IGT and/or IFG were similar across different clusters, even for those with higher risk of T2D during follow-up (i.e., Cluster 5 and Cluster 6) which did not show worse glycemic status at baseline compared to other clusters. These findings imply that our genetic subtyping approach might help identify individuals with high risk of future T2D risk before they manifest obvious glycemic change during T2D development, and also reflect the advantage of using the stable genetic information compared to T2D-related metabolic phenotypes which are subject to change with disease progression or treatment.^11^ Genetic-based subtyping of prediabetes may have important implications in early prevention of T2D, since previous phenotype-based studies identified high T2D risk subgroups which already showed worse glycemic status at baseline compared to low T2D risk subgroups.^4^ It is worthwhile to mention that clusters of prediabetes identified in this study showed no significant differences in demographic, socioeconomic, and behavioral factors in both cohorts, further supporting the robustness of results using genetic information with minimum confounding issues.

It is intriguing that individuals with prediabetes in Cluster 6 showed the highest risk of T2D since this cluster had a very low proinsulin genetic score which was associated with a favorable metabolic profile (e.g., high HOMA-B and low levels of glycemic traits) and reduced risk of T2D in the current and previous studies.^11^ However, low proinsulin genetic score was associated with high fasting proinsulin levels adjusted for fasting insulin.^11^ The proinsulin to insulin ratio has been used to differentiate disproportionally elevated proinsulin from compensatory hyperinsulinemia,^30^ and a high proinsulin to insulin ratio might indicate disturbed insulin secretion^31-33^ and has been associated with increased risk ofT2D.^30 34 35^ Thus, high risk of T2D might be partially explained by disturbed insulin secretion among individuals with prediabetes in this cluster. Our further analysis showed a potential interaction between the proinsulin score and other genetic scores, suggesting that the genetic effects of other biological pathways on risk of T2D might be strengthened among individuals with a low proinsulin score indicating disturbed insulin secretion. This might help explain high risk of T2D for individuals with prediabetes in Cluster 6, since levels of the other four genetic scores were slightly above average in this cluster. However, the underlying biological mechanisms are unclear and need to be clarified.

This study identified another potential genetic subtype of prediabetes with high T2D risk, Cluster 5, which was characterized by high levels of lipodystrophy-like, liver/lipid and proinsulin scores but low levels of beta-cell function and obesity score. The metabolic trait profile of this cluster was also mixed. It had the lowest levels of HDL-C and highest levels of HOMA-IR among all clusters, but also some favorable levels of metabolic traits (e.g., high HOMA-B and low-BMI). Individuals with prediabetes in this cluster might share some features with previously suggested normal-weight “metabolically obese”,^36^ and their elevated risk of T2D might be due to lipodystrophy or fat distribution–related (e.g., reduced subcutaneous adiposity) insulin resistance.^11 36^ However, lipodystrophy or fat distribution was not measured in this study, and future studies with these measures might help explain the elevated risk of T2D in this cluster of individuals with prediabetes.

Another major finding of this study is that adherence to healthy lifestyle was associated with decreased risk of T2D across different clusters of individuals with prediabetes, except for Cluster 3. This cluster was only identified in HCHS/SOL (Hispanics/Latino adults in the US) but not in UKBB (non-Hispanic whites), and the non-significant association between the health lifestyle score and risk of T2D in this cluster might be due to a relatively small sample size. Interestingly, we identified a cluster of individuals with prediabetes, Cluster 5, which might experience greater benefits from adhering to a healthy lifestyle in prevention of T2D. Individuals with prediabetes in Cluster 5 had ∼30% greater risk of T2D compared to those in Cluster 1, but adherence to a healthy lifestyle lowered the risk of T2D to a similar level for both these clusters. Our study further emphasizes the importance of healthy lifestyle for all individuals with prediabetes in reducing their risk of progression to diabetes,^37 38^ and suggests a potential genetic subtype of prediabetes that could have extra benefits of healthy lifestyle. This may have important public health implications.

### Strengths and limitations of the study

Our study has several strengths. We used data from two prospective cohorts with large sample size of participants with prediabetes, especially in UKBB, which help to reduce inconsistency of clustering results among multiple studies caused by small sample size and reproducibly subtype prediabetes into five clusters. Our subtyping approach only used genetic information, which indicates that subtyping result will not change over time. Longitudinal data helped to examine the risk of incident diabetes across clusters to point out the advantage of this approach. With well-collected lifestyle factors, we were able to estimate health lifestyle score to evaluate the benefit of adherence to health lifestyle across clusters.

This study has several limitations. Both cohorts lacked data on some metabolic traits, such as blood proinsulin levels and fat distribution measures, which can help better understand the underlying mechanisms for genetic subtypes of prediabetes with high risk of T2D. Although our subtyping approach was based on multiple T2D-assocaited genetic variants and five different pathways related to T2D, these genetic variants might only explain a small portion of genetic risk for T2D. More studies are needed to reveal genetic architecture of T2D and related quantitative traits, which will yield more biological pathways related to T2D and thus help identify more accurate subtypes of prediabetes based on genetic information. This study included US Hispanics/Latino and non-Hispanic white individuals. Although findings were consistent between these two study populations, further investigations in other racial/ethnic groups are needed given the heterogeneity in genetics of T2D across different populations.^39^

## Conclusions

In this analysis of US Hispanics/Latinos from HCHS/SOL and non-Hispanic white individuals from UKBB, we identified several clusters of prediabetes based on genetic information, and two potential genetic subtypes of prediabetes showed relatively high risk of T2D over time. We also observed generally favorable relationships between healthy lifestyle and risk of T2D among individuals with prediabetes, regardless of their genetic subtypes, though individuals in one subtype may realize extra benefits in terms of risk reduction from a healthy lifestyle. This study extends the application of genetic information in subtyping of prediabetes and provides useful information to early prevention and intervention among people with high T2D risk.

## Data Availability

All data produced in the present work are contained in the manuscript

## Contributors

All authors were involved in conceiving and designing the study. YL did the statistical analyses and wrote the manuscript. All authors interpreted the data, and critically revised the manuscript. All authors approved the final version of the manuscript and agree to be accountable for the accuracy of the work. QQ supervised the study and is the guarantor. The corresponding author attests that all listed authors meet authorship criteria and that no others meeting the criteria have been omitted.

The authors thank Dr. Miriam S. Udler for her suggestion in constructing partitioned polygenetic risk scores. The authors thank the staff and participants of HCHS/SOL for their important contributions. A complete list of HCHS/SOL staff and investigators can be found in Ann Epidemiol 2010; 20:642–649 or at http://sites.cscc.unc.edu/hchs/.

## Ethical approval

HCHS/SOL was approved by the institutional review boards at Albert Einstein College of Medicine. UKBB has approval from the North West Multi-centre Research Ethics Committee (MREC) as a Research Tissue Bank (RTB) approval. Informed consent was implied with the return of the self-administered questionnaires.

