## Supplementary materials for "Genetic subtypes of prediabetes, healthy lifestyle, and risk of type 2 diabetes: Prospective cohort study"

### Supplementary Methods

### Study population

HCHS/SOL has 16,415 participants aged 18-74 years at the screening from four U.S. metropolitan areas (Bronx, NY; Chicago, IL; Miami, FL; and San Diego, CA) during 2008-2010.**^1 2^** In terms of Hispanic/Latino background, the Bronx field center sample is predominantly Puerto Rican and Dominican, while the majority of participants in the San Diego site are Mexican in origin. Study participants in the Miami field center are Cuban and Central/South American, and participants in the Chicago field center are Mexican, Puerto-Rican, and Central/South American. A minimum of 2,000 participants in each of the pre-specified Hispanic/Latino groups (Mexican, Puerto Rican, Cuban, and Central/South American) is required to support the analysis objectives, and sample sizes are monitored continuously to determine if adjustments to the sampling strategy are needed. A comprehensive battery of interviews relating to personal and family characteristics and health status and behaviors, as well as a clinical assessment with blood draw, were conducted at an in-person clinic baseline visit. Each participant accepted a clinical examination, including clinical assessments, questionnaires, and anthropometric measurements. Standardized instruments were used to determine information about health behaviors, medical history, demographics, education, socioeconomic status, and diet. Participants were required to fast for at least 8 h prior to the visit, consuming only water and necessary medications. Throughout the 3-year exam period, collection of blinded repeat blood samples and repeated random measurements of clinical procedures were used as quality control procedures. Participants are contacted by telephone approximately 6 weeks after the baseline visit to obtain a second 24-hour dietary recall. The analyses described here included 3,550 participants with normal glucose and 3,677 participants with prediabetes (**Supplementary Table 3**). The study was approved by each of the Field Center’s and the Coordinating Center’s Institutional Review Board. All enrollment participants provided signed informed consent.

UKBB is a large prospective observational study that recruited approximately 500,000 men and women of various ethnicities aged 37-73 years from across 22 centers located throughout England, Wales, and Scotland between 2006 and 2010.^3^ UKBB study design and population have been described in detail previously.^3^ In brief, UKBB study recruited more than 500,000 participants aged 37 to 73 from 2006 to 2010. Participants underwent various measurements including a standardized baseline laboratory assessment and provided a wide range of information on health and diseases at recruitment. Information on lifestyle and other health-related aspects were collected through baseline questionnaires, interviews, and physical measurements. Blood samples were collected for genotyping. To date, UKBB has completed up to four times by over 200,000 participants, and from the first repeat of the entire baseline assessment in around 20,000 participants. UKBB received ethical approval from the research ethics committee (REC reference for UKBB 11/NW/0382), and participants provided written informed consent. The current study was conducted under application number 56483 of UKBB resource. In the current analysis, 400,149 non-Hispanic white participants without diabetes at baseline and had genetic data were included, including 383,865 participants with normal glucose and 16,284 participants with prediabetes (**Supplementary Table 4**).

### Ascertainment of prediabetes and T2D

In HCHS/SOL, defining prediabetes and diabetes was based on the criteria of American Diabetes Association. Glycemic indices, including fasting plasma glucose (fasting glucose), 2-hour oral glucose tolerance test (OGTT 2h glucose) and hemoglobin A1c (HbA1c), and fasting plasma insulin were measured to define prediabetes and diabetes. Participants with prediabetes were defined as those with fasting glucose between 100-125 mg/dL, OGTT 2h glucose between 140-199 mg/dL, or HbA1c between 5.7% and 6.5%.**^4^** Participants with diabetes were defined as those with fasting glucose levels ≥126 mg/dL, 2h glucose after OGTT ≥200 mg/dL, HbA1c ≥6.5%, or current use of anti-diabetic medications.**^4^** Participants free of diabetes at baseline (Visit 1) who were identified as having diabetes at Visit 2 were deemed to be incident cases of T2D.

In the UK Biobank, HbA1c were used as the main glycemic index to define prediabetes and T2D. Prevalent diabetes at baseline was identified through multiple procedures and sources of the diagnosis (e.g., self-report and medical record)**^5^** in addition to potential undiagnosed diabetes (HbA1c ≥6.5% [48 mmol/mol]).**^4^** Participants with prediabetes were defined as those with HbA1c between 5.7% and 6.5%.**^4^** Diabetes that occurred during follow-up was identified using primary care, hospital admissions, and self-reported, and incident T2D was defined by ICD-10 code E11 in Field 130708 and 130709.**^5 6^** The data fields have been generated by mapping: Read code information in the Primary Care data (Category 3000), ICD-9 and ICD-10 codes in the Hospital inpatient data (Category 2000), ICD-10 codes in Death Register records (Field 40001, Field 40002), and Self-reported medical condition codes (Field 20002) reported at the baseline or subsequent UK Biobank assessment center visit to 3-character ICD-10 codes (biobank.ndph.ox.ac.uk/showcase/label.cgi?id=1712)**.** Participants with HbA1c ≥6.5% were also identified as diabetes.**^5^**

### Measurements of metabolic traits

In HCHS/SOL, BMI was calculated as weight in kilograms divided by height in meters squared. Waist-to-hip ratio (WHR) is the ratio of the circumference of the waist to that of the hips. Centralized laboratory tests were performed to determine plasma fasting glucose, 2h glucose after OGTT, fasting insulin, and hemoglobin A1c (HbA1c), and serum lipids including triglycerides, and total, low-density lipoprotein (LDL), and high-density lipoprotein (HDL) cholesterol. Homeostatic model assessment of insulin resistance (HOMA-IR) and beta-cell function (HOMA-B) were derived using equations based on fasting glucose and insulin.^7^ In the UK Biobank, metabolic traits included BMI, WHR, random glucose, HbA1c, triglycerides, and total-, LDL- and HDL-cholesterol.^8^

### Genotyping, pPRSs calculation and clustering

In HCHS/SOL, genome-wide genotyping was performed in 12,633 participants using a customized Illumina array (15041502 B3; llmina Omni 2.5M array plus ~150K custom SNPs, and imputation was carried out based on the 1000 Genomes Project phase 3 reference panel.**^9^** DNA extracted from blood was genotyped on an Illumina custom array, SOL HCHS Custom 15041502 B3, consisting of the Illumina Omni 2.5M array (HumanOmni2.5-8v1-1) and ~150,000 custom SNPs selected to include ancestry-informative markers, variants characteristic of Amerindian populations. Samples were checked for annotated sex or genetically determined sex, gross chromosomal nomalies,42 unexpected duplicates, missing call rates, contamination, and batch effects. A total of 12,803 samples passed quality control with a missing call rate <1%. Quality metrics used to filter SNPs for the imputation basis and association testing included missing call rate (>2%), Mendelian errors, duplicate-sample discordance, and deviation from Hardy-Weinberg equilibrium (p<10e5 in a meta-analysis of nine groups within which individuals had both parents from the same country of origin). A total of 2,232,944 SNPs passed quality metrics and were informative. Genome-wide imputation was carried out using the 1000 Genomes Project phase 1 reference panel SHAPEIT2 and IMPUTE2 software, as described previously.^9^ Five pPRSs, beta-cell score, proinsulin score, obesity score, lipodystrophy-like score and liver-lipid scores, were calculated based on 94 T2D-assocaited genetic variants using a previously reported method.**^10^** According to the study, 94 variants were partitioned into five SNP sets in Bayesian nonnegative matrix factorization (bNMF) with the partitioning process being supervised by 47 T2D-related traits. Within each SNP sets, a polygenetic risk score was assessed by computing the sum of risk alleles of SNPs in the set for each individual, weighted by their effect sizes estimated from bNMF.

In the UK Biobank, genome-wide genotyping was performed in ~500,000 participants using Affymetrix UK BiLEVE Axiom Array or Affymetrix UK Biobank Axiom Array.**^11^** The genotyping process and quality control used in UKBB has been described elsewhere in more details.^11^ UKBB estimated haplotypes for the full cohort (pre-phasing), followed by haploid imputation. For the pre-phasing step, only markers present on both the UK BiLEVE and UK Biobank Axiom arrays were used. Markers that failed quality control in more than one batch, had a greater than 5% overall missing rate, and had a MAF of less than 0.0001 were removed. Samples that were identified as outliers for heterozygosity and missing rate were removed. These filters resulted in a dataset with 670,739 autosomal markers in 487,442 samples. The 1000 Genomes phase 3 dataset was used as a reference panel, predominantly to help with the phasing of samples with non-European ancestry. Haplotype Reference Consortium (HRC) data was used as the main imputation reference panel because it consisted of the largest available set (64,976) of broadly European haplotypes at 39,235,157 SNPs. We only considered participants of white British descent in the current study.

K-means consensus clustering was performed in 3,677 participants with prediabetes in HCHS/SOL and 16,284 participants with prediabetes in the UK Biobank, respectively, based on the five pPRSs as input features, using the ConsensusClusterPlus R package**.^12^** A subset of participants were selected, and Pearson’s correlation coefficient among them was calculated using five pPRSs. Then, K-means clustering was performed based on the correlation matrix, with each participant being assigned into one cluster. The whole process was repeated 1,000 times. The cluster memberships of the single clustering result were combined to a consensus matrix, determining the number of times each pair of samples was clustered together over all clusterings. The following settings were used for clustering: number of repetitions = 1,000 bootstraps; pItem = 0.8 (resampling 80% of any sample); pFeature = 1.0 (using all pPRSs). The number for whole clusters was set from two to 12 clusters. The best number of clusters was determined by the largest average Silhouette coefficients. We selected a 6-cluster for HCHS/SOL as it had highest average Silhouette coefficient of 0.371, and a 5-cluster for UBKBB with the highest average Silhouette coefficient of 0.381 (**Supplementary Figure 4**). Based on the evidence above, prediabetic participants in HCHS/SOL were classified into six subgroups, and those in UKBB were classified into five subgroups (**Figure 1**).

### Lifestyle score calculation

Adherence to a healthy lifestyle was measured by a lifestyle score based on five well-established modifiable factors including BMI, smoking, alcohol drinking, physical activity, and diet for T2D.**^13^** Each factor was assigned with a score, and then an overall lifestyle score was calculated by summing up five scores (**Supplementary Table 1 and 2**). In HCHS/SOL, both smoking and drinking status was measured by self-report and categorized into three categories: current, former, and never. If participants reported current alcohol drinking, they were asked to provide the number of drinks they consumed in a week. Physical activity was measured using the Global Physical Activity Questionnaire.**^14^** Dietary intake was estimated using the National Cancer Institute methodology based on two 24-h dietary recalls and a food propensity questionnaire.**^15^** Overall dietary quality was estimated by the Alternative Healthy Eating Index (AHEI)-2010.**^16^** Each factor was assigned with a continuous score. The participants received 2 points if 18.5≤ BMI ≤24.9 kg/m2, or 1 point if 25≤ BMI ≤29.9 kg/m2, or 0 if BMI ≥30 kg/m2. AHEI2010 was used to present dietary quality and values within four quartiles were given 0, 1, 2 and 3 correspondingly. The participants were assigned with 2 if he or she is never smoker, 1 if ever smoker or 0 if current smoker. Alcohol use level was a binary score: current alcohol use <7 drinks/week was given 1 point for females and <14 drinks/week for males. Total physical activity (MET-min/day) from Global physical activity questionnaire (GPAQ) were cut into quartiles with each quartile being assigned with 0, 1, 2 and 3 points. It should be noted that AHEI2010 quartiles and physical activity quartiles were calculated for males and females separately. Finally, five scores were summed up as healthy lifestyle score.

In the UK Biobank, similar approaches were used to measure lifestyle factors and calculate the lifestyle score (**Supplementary Table 2**). Since the AHEI-2010 was not available in the UK Biobank, we assessed overall dietary quality based on 5 major food groups, as described in Supplementary Table 2. Five food groups, including Red and processed meat, Fresh fruit, Whole grain, Refined grain and Fish, were selected according to previous studies,^17-19^ and each food group was given one score or zero based on their serving per day. Five scores were summed up as healthy lifestyle score.

In the current analysis, participants with the 2^nd^ and 3^rd^ tertiles of the lifestyle score were defined as adherence to a healthy lifestyle.

### Statistical Analysis

After calculating pPRSs, linear regression was used to examine associations of five pPRSs with baseline metabolic traits after adjustment for covariates in both cohorts. To examine the association between five pPRSs and risk of T2D (per SD increment) among participants without diabetes at baseline, Robust Poisson regression adjusting for covariates was performed in HCHS/SOL, while Cox proportional-hazards regression adjusting for covariates was used in UKBB.

After obtaining clusters of prediabetes, Analysis of Variance (ANOVA) and Chi-squared tests were applied to test differences in five pPRSs and characteristics including continuous variables (e.g., baseline glycemic traits and changes in glycemic traits) and categorical variables (e.g., glycemic status [impaired glucose tolerance (IGT), only impaired fasting glucose (IFG), or both] and Hispanic/Latino background) across clusters of prediabetes in both cohorts. To compare the relative risks of T2D across clusters of prediabetes (with Cluster 1 defined as reference), Robust Poisson regression and Cox proportional-hazards regression adjusting for covariates were performed in HCHS/SOL and UKBB, respectively.

To detect the interaction between proinsulin score and the other four pPRSs and a summed score (summing up these four pPRSs), Robust Poisson regression and Cox proportional-hazards regression adjusting for covariates were performed after participants without diabetes were stratified by the median of the proinsulin score (interactions were test by including the respective interaction terms in the models) in HCHS/SOL and UKBB, respectively.

The associations between adherence to a healthy lifestyle and risk of T2D in all participants with prediabetes, as well as across clusters of prediabetes, were estimated using Robust Poisson regression and Cox proportional-hazards regression adjusting for covariates in HCHS/SOL and UKBB, respectively. The relative risks of T2D comparing Cluster 5 to Cluster 1 while stratifying according to the lifestyle score (with Cluster 1 with unhealthy lifestyle defined as reference) were also assessed in both cohorts using Robust Poisson regression and Cox proportional-hazards regression adjusting for covariates, respectively.

For all the analyses above, RRs and 95% confidence intervals were reported when using Robust Poisson regression in HCHS/SOL, and HRs and 95% confidence intervals were reported when using Cox proportional-hazards regression in UKBB. Results from HCHS/SOL and UKBB were pooled by inverse-variance weighted, fixed-effects meta-analyses.

In the combined analysis, difference in the effect sizes of healthy lifestyle on risk of T2D between clusters was tested by Cochran's Q test for heterogeneity. We considered 2-sided P values <0.05 as statistically significant and used False Discovery Rate adjusted P value for multiple tests. All analyses were performed using R 4.1.2.

### Supplementary Figure 1 Distribution of five pPRSs among participants without diabetes at baseline in HCHS/SOL and in UKBB

Histogram of Beta cell score, Proinsulin score, Obesity score, Lipodystrophy-like score and Liver-lipid score among participants without diabetes at baseline in HCHS/SOL **(A)** and UKBB **(B)**.

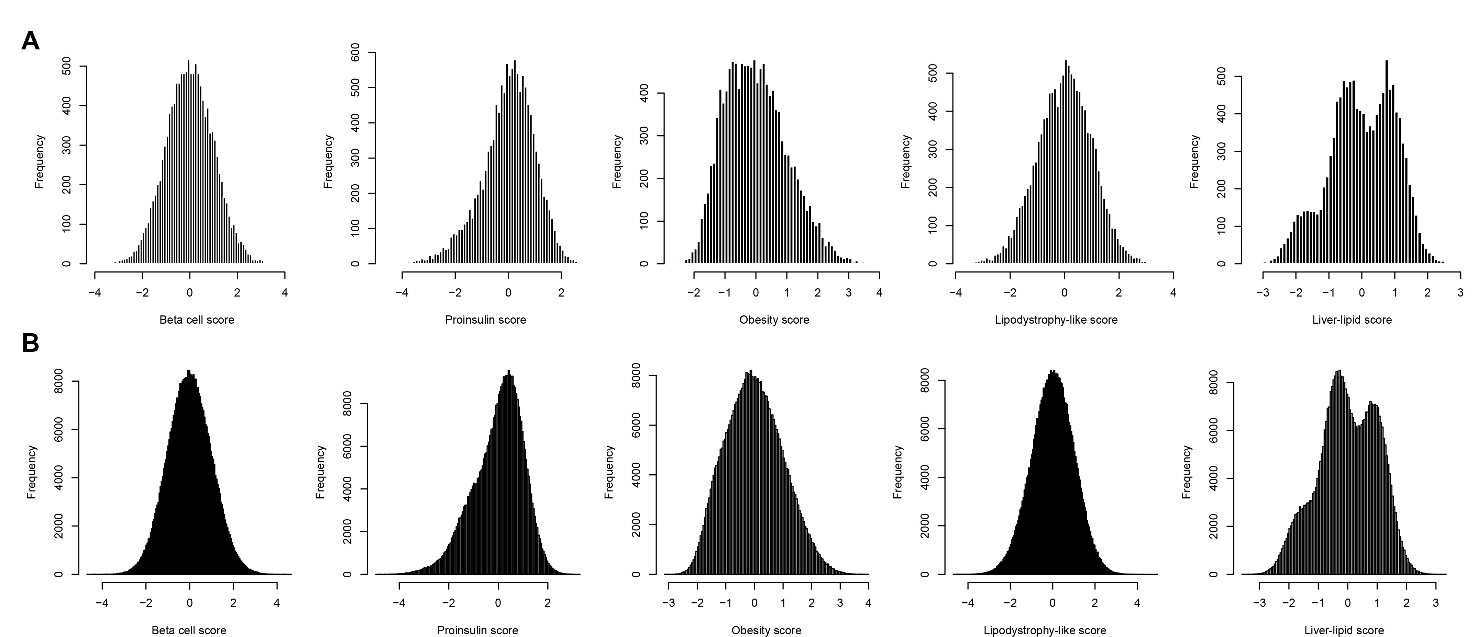

### Supplementary Figure 2 Pearson’s correlation coefficients among five pPRSs in HCHS/SOL and in UKBB

Pearson’s correlation coefficients among Beta cell score, Proinsulin score, Obesity score, Lipodystrophy-like score and Liver-lipid score among participants without diabetes at baseline in HCHS/SOL **(A)** and UKBB **(B)**.

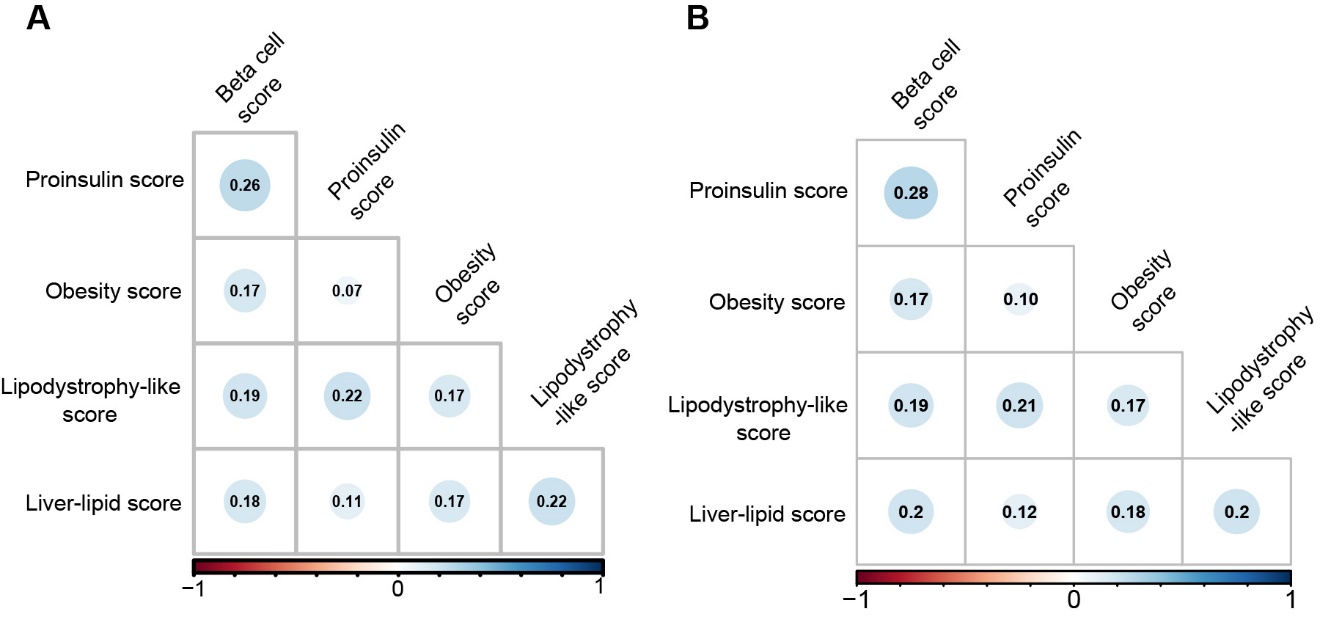

### Supplementary Figure 3 Associations of pPRSs with T2D-related metabolic traits and incident T2D in HCHS/SOL and in UBKK

**(A)** Heatmap of the associations between pRPSs and T2D-related metabolic traits among participants without diabetes at baseline in HCHS/SOL. Data are beta coefficients estimated by linear regression models after adjustment for age, sex, U.S.-born status, Hispanic/Latino background, smoking, drinking, education, annual income, eigenvectors derived from GWAS, and lipid-lowering medicine use (only for TC, TG, LDL and HDL). **(B)** Heatmap of the associations between pRPSs and T2D-related metabolic traits among participants without diabetes at baseline in UKBB. Data are beta coefficients estimated by linear regression models after adjustment for age, sex, smoking, drinking, education, Townsend deprivation score, eigenvectors derived from GWAS, and lipid-lowering medicine use (only for TC, TG, LDL and HDL). **(C)** Associations between pPRSs and incident T2D among participants without diabetes at baseline in HCHS/SOL. Data are risk ratios (RRs) and 95% confident intervals (CIs) estimated by Poisson regression after adjustment for age, sex, U.S.-born status, Hispanic/Latino background, education, annual income, eigenvectors derived from GWAS. **(D)** Association between pPRSs and incident T2D among participants without diabetes at baseline in UKBB. Data are Hazard ratios (HRs) and 95% CIs estimated by Cox proportional regression after adjustment for age, sex, education, Townsend deprivation score and eigenvectors derived from GWAS.

Abbreviations: HOMA-B, Homeostatic Model Assessment for Beta Function; HOMA-IR, Homeostatic Model Assessment for Insulin Resistance; OGTT, Oral Glucose Tolerance Test; HbA1C, Hemoglobin A1C; BMI, Body Mass Index; TC, Total Cholesterol; LDL, Low-density Lipoprotein Cholesterol; HDL, High-density Lipoprotein Cholesterol; TG, Triglyceride.

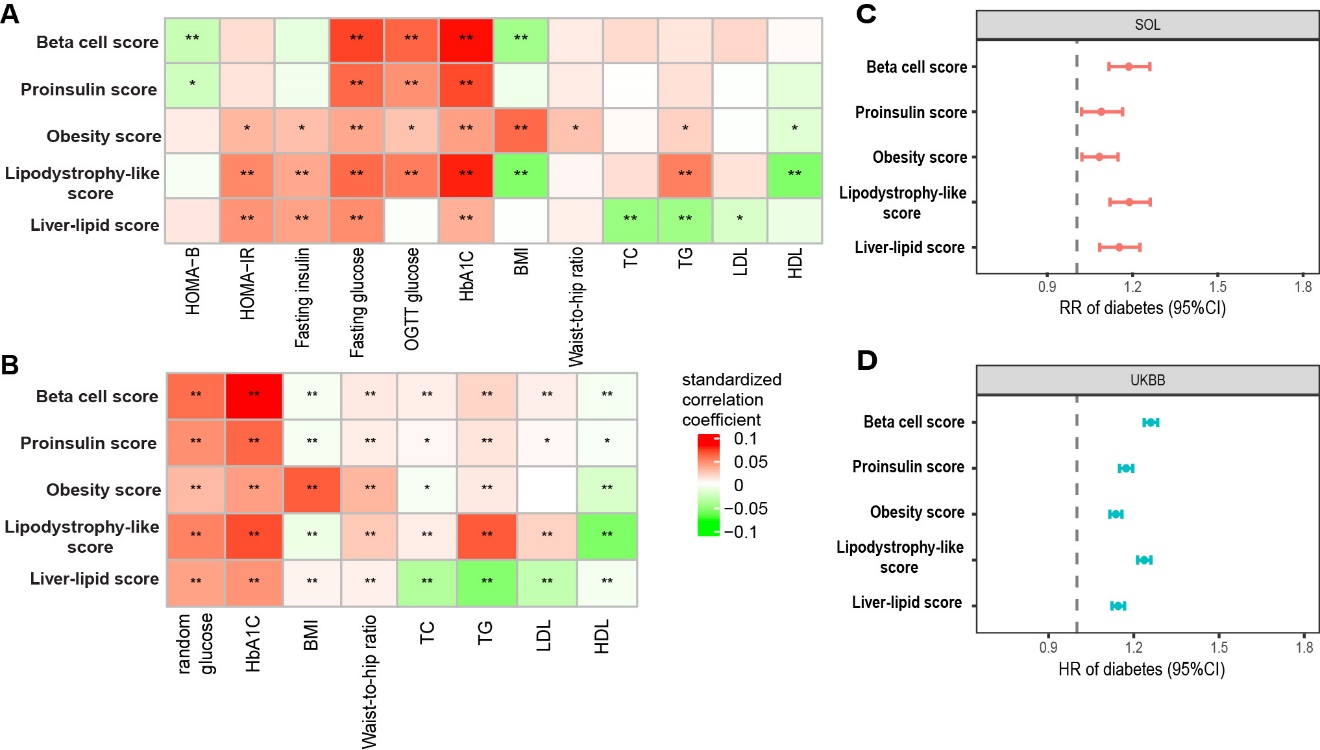

### Supplementary Figure 4 Silhouette analysis in K-means-based consensus clustering

**(A)** Silhouette coefficients at different number (k) of clusters in HCHS/SOL and in UKBB. (B) Silhouette coefficients for each participant in each cluster when k=6 in HCHS/SOL. (C) Silhouette coefficients for each participant in each cluster when k=5 in UKBB.

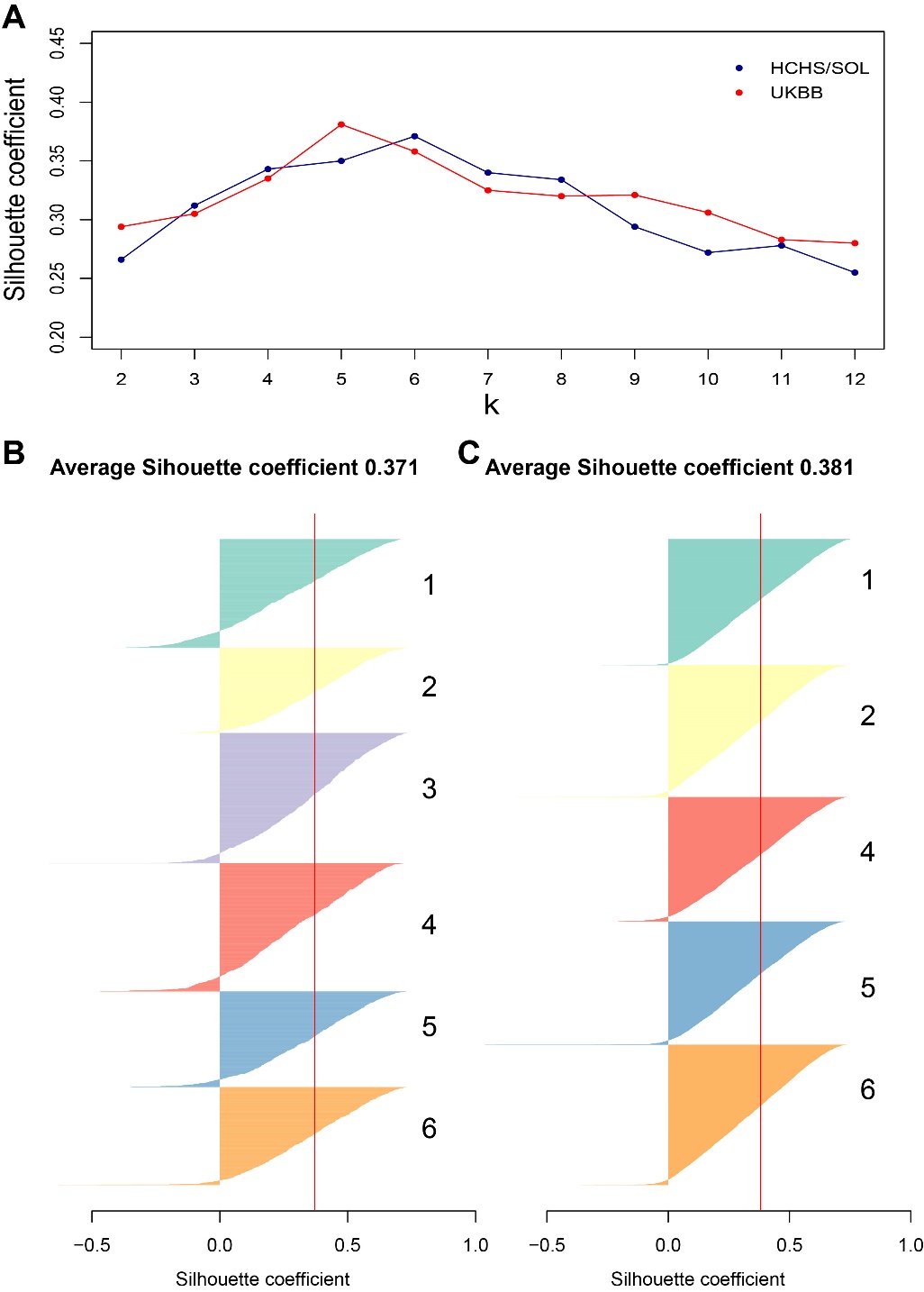

### Supplementary Figure 5 Comparison of five pRPSs and five T2D-related metabolic traits among clusters of prediabetes

Circos plot shows comparison of five pPRSs (red links) and five T2D-related metabolic traits (blue links) between any two clusters of individuals with prediabetes in HCHS/SOL (only comparison with P<0.05 after FDR are presented with links). Numbers in the parentheses indicate numbers of significant differences (lower/higher) in the corresponding variable between two clusters. Arrows of links point to the cluster with a higher value of the corresponding variable compared to other clusters.

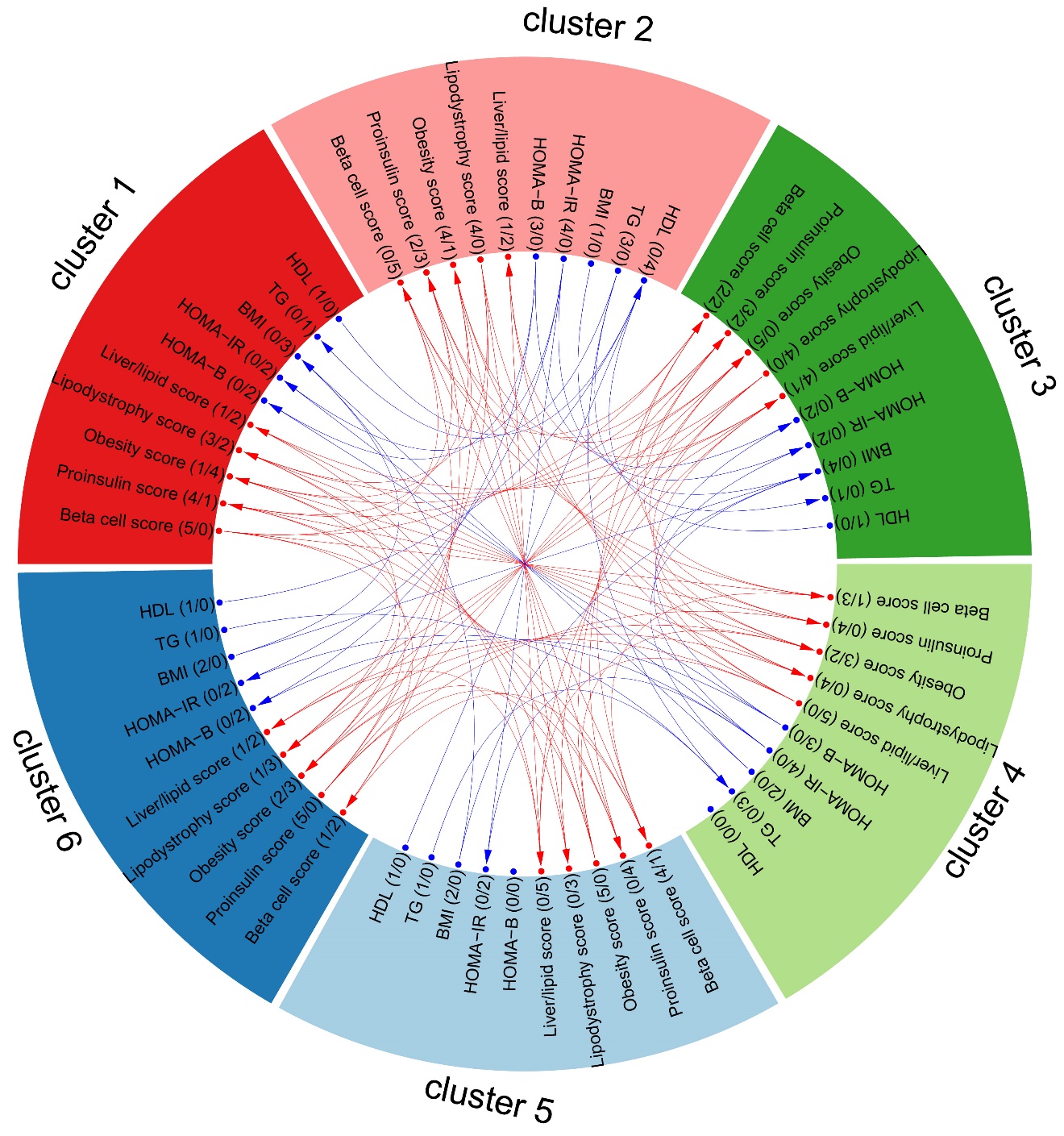

### Supplementary Figure 6 Proportion of six clusters across Hispanic/Latino groups in HCHS/SOL

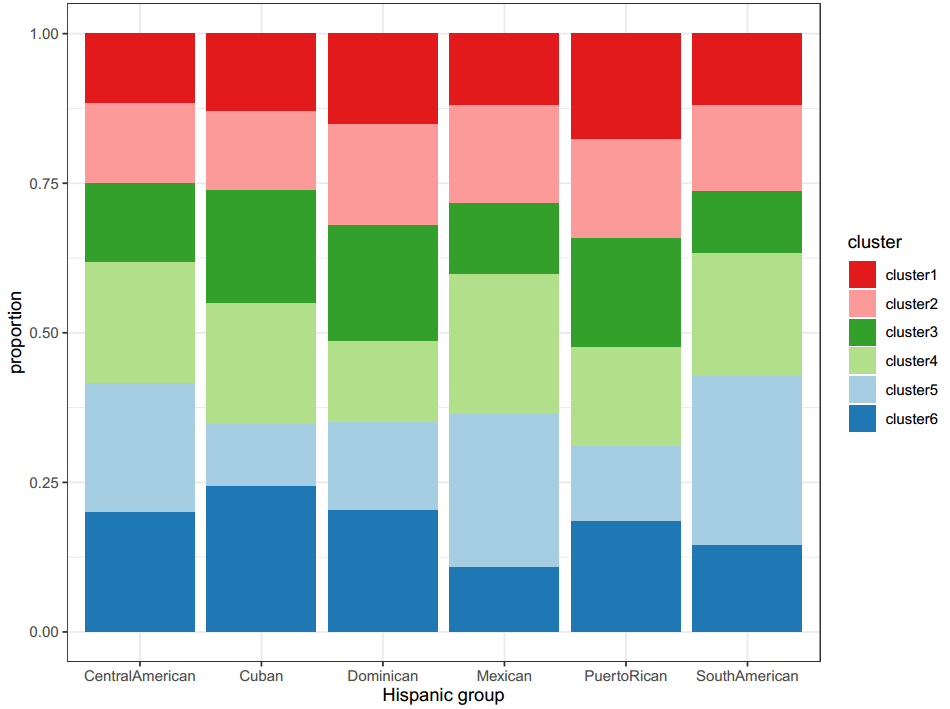

### Supplementary Table 1 Components of healthy lifestyle score in HCHS/SOL

| Lifestyle factors | Category | Score |
| --- | --- | --- |
| BMI | 18.5–24.9 kg/m2 | 2 |
|  | 25-29.9 kg/m2 | 1 |
|  | ≥30 kg/m2 | 0 |
| AHEI-2010 | AHEI Q1 | 0 |
| (sex specific) | AHEI Q2 | 1 |
|  | AHEI Q3 | 2 |
|  | AHEI Q4 | 3 |
| Smoking | Current | 0 |
|  | Ever | 1 |
|  | Never | 2 |
| Alcohol use level | Female: current use <7 drinks/week Male: current use <14 drinks/week | 1 |
|  | other | 0 |
| Physical activity | Total MET Q1 | 0 |
| (sex specific) | Total MET Q2 | 1 |
|  | Total MET Q3 | 2 |
|  | Total MET Q4 | 3 |

AHEI-2010, Alternate Healthy Eating Index 2010; MET, Metabolic Equivalent of Task; BMI, Body Mass Index; Q1, the first quartile; Q2, the second quartile; Q3, the third quartile; Q4, the fourth quartile.

### Supplementary Table 2 Components of healthy lifestyle score in UKBB

| Lifestyle factors | Category | Score |
| --- | --- | --- |
| BMI | ≤18.5 kg/m2 | 0 |
|  | 18.5–24.9 kg/m2 | 2 |
|  | 25-29.9 kg/m2 | 1 |
|  | ≥30 kg/m2 | 0 |
| Diet | Number of the following:  1. Red and processed meat <3.5 serving/week AND processed meat <1 serving/week  2. Fresh fruit ≥ 3 serving/day OR fresh vegetable ≥ 3 serving/day OR a combination ≥ 4.5 serving/day  3. Whole grain ≥ 3 serving/day  4. Refined grain < 1.5 serving/day  5. Fish ≥ 2 serving/day | 0-5 |
| Smoking | Current | 0 |
|  | Ever | 1 |
|  | Never | 2 |
| Alcohol use level | Female: current use <7 drinks/week Male: current use <14 drinks/week | 1 |
| Physical activity | Total MET Q1 | 0 |
| (sex specific) | Total MET Q2 | 1 |
|  | Total MET Q3 | 2 |
|  | Total MET Q4 | 3 |

BMI, Body Mass Index; MET, Metabolic Equivalent of Task; Q1, the first quartile; Q2, the second quartile; Q3, the third quartile; Q4, the fourth quartile;

### Supplementary Table 3 Baseline characteristics of participants with normal glucose and participants with prediabetes in HCHS/SOL

|  | | Normal glucose | Prediabetes | *P* value | |
| --- | --- | --- | --- | --- | --- |
| Number | 3,550 | | 3,677 | |  |
| Age (years) | 39.0(27.0-49.0) | | 50.0(42.0-57.0) | | <0.001 |
| Sex |  | |  | |  |
| Female | 4184(61.0) | | 3648(57.7) | | <0.001 |
| Male | 2672(39.0) | | 2672(42.3) | |  |
| U.S.-born status |  | |  | |  |
| non U.S.-born | 5197(76.0) | | 5436(86.2) | | <0.001 |
| U.S.-born | 1645(24.0) | | 874(13.8) | |  |
| Smoking status |  | |  | |  |
| Never | 4374(64.1) | | 3750(59.4) | | 0.046 |
| Former | 1043(15.2) | | 1325(21.0) | |  |
| Current | 1412(20.7) | | 1235(19.6) | |  |
| Drinking status |  | |  | |  |
| Never | 3247(47.4) | | 3284(52.0) | | 0.122 |
| Former | 3197(46.8) | | 2707(42.9) | |  |
| Current | 400(5.8) | | 319(5.1) | |  |
| Hispanic/Latino background |  | |  | |  |
| Central American | 588(11.3) | | 556(11.2) | | 0.013 |
| Cuban | 917(17.6) | | 941(19.0) | |  |
| Dominican | 523(10.1) | | 434(8.8) | |  |
| Mexican | 1887(36.3) | | 1833(37.0) | |  |
| Puerto Rican | 844(16.2) | | 833(16.8) | |  |
| South American | 447(8.6) | | 356(7.2) | |  |
| Education |  | |  | |  |
| Less Than High School | 2070(30.3) | | 2556(40.6) | | <0.001 |
| High School or Equivalent | 1965(28.8) | | 1548(24.6) | |  |
| Greater than High School or Equivalent | 2803(41.0) | | 2198(34.9) | |  |
| Annual income |  | |  | |  |
| <$10,000 | 859(13.7) | | 906(15.6) | | 0.723 |
| $10,001-$20,000 | 1969(31.6) | | 1871(32.2) | |  |
| $20,001-$40,000 | 2181(35.0) | | 1989(34.2) | |  |
| $40,001-$75,000 | 914(14.7) | | 796(13.7) | |  |
| >$75,000 | 304(4.9) | | 252(4.3) | |  |
| AHEI-2010 | 46.7(41.9, 52.3) | | 49.2(44.2, 54.8) | | 0.655 |
| Total physical activity (MET-min/day) | 257.1(51.4, 960.0) | | 177.1(22.9, 788.6) | | 0.317 |
| BMI (kg/m^2^) | 27.3(24.3, 30.9) | | 29.7(26.6, 33.4) | | <0.001 |
| Waist-to-hip ratio | 0.90(0.85, 0.95) | | 0.93(0.89, 0.97) | | <0.001 |
| HOMA-Β | 117.8(77.1, 175.2) | | 121.9(80.4, 184.8) | | <0.001 |
| HOMA-IR | 1.9(1.2, 2.9) | | 2.8(1.8, 4.3) | | <0.001 |
| Fasting insulin (mU/L) | 8.6(5.6, 12.7) | | 11.6(7.6, 17.6) | | <0.001 |
| Fasting glucose (mg/dL) | 90.0(86.0, 94.0) | | 99.0(92.0, 104.0) | | <0.001 |
| OGTT 2h glucose (mg/dL) | 101.0(86.0, 116.0) | | 133.0(108.0, 155.0) | | <0.001 |
| HbA1C (%) | 5.3(5.1, 5.5) | | 5.7(5.5, 5.9) | | <0.001 |
| TC (mg/dL) | 187.0(161.0, 213.0) | | 205.0(178.0, 232.0) | | <0.001 |
| LDL (mg/dL) | 113.0(92.0, 136.5) | | 127.0(104.0, 152.0) | | <0.001 |
| HDL (mg/dL) | 49.0(42.0, 58.0) | | 47.0(40.0, 56.0) | | <0.001 |
| TG (mg/dL) | 94.0(66.0, 137.0) | | 125.00(90.0, 177.6) | | <0.001 |

Values are presented as median (IQR) for continuous variables or N (%) for categorical variables. Continuous variables were adjusted for age and sex in multiple linear regression model when compare their difference between participants with normal glucose and those with prediabetes. Participants who took lipid-lowering medication were excluded when analyzing TC, TG, LDL and HDL.

Abbreviations: AHEI-2010, Alternate Healthy Eating Index 2010; MET, Metabolic Equivalent of Task; BMI, Body Mass Index; HOMA-B, Homeostatic Model Assessment for Beta Function; HOMA-IR, Homeostatic Model Assessment for Insulin Resistance; OGTT, Oral Glucose Tolerance Test; HbA1C, Hemoglobin A1C; TC, Total Cholesterol; LDL, Low-density Lipoprotein Cholesterol; HDL, High-density Lipoprotein Cholesterol; TG, Triglyceride.

### Supplementary Table 4 Baseline characteristics of participants with normal glucose and participants with prediabetes in UKBB

|  | | Normal glucose | Prediabetes | *P* value | |
| --- | --- | --- | --- | --- | --- |
| Number | 383,865 | | 16,284 | |  |
| Age (year) | 58.0(50.0-63.0) | | 62.0(57.0-66.0) | | <0.001 |
| Sex |  | |  | |  |
| Female | 211,555(55.1) | | 8,209(50.4) | | <0.001 |
| Male | 172,310(44.9) | | 8,075(49.6) | |  |
| Smoking status |  | |  | |  |
| Never | 213,166(55.7) | | 7,442(45.9) | | <0.001 |
| Former | 131,148(34.3) | | 6,220(38.4) | |  |
| Current | 38,226(10.0) | | 2,538(15.7) | |  |
| Drinking status |  | |  | |  |
| Never | 12,661(3.3) | | 1,019(6.3) | | <0.001 |
| Former | 12,282(3.2) | | 8,46(5.2) | |  |
| Current | 358,577(93.5) | | 14,392(88.5) | |  |
| Education |  | |  | |  |
| College or University degree | 122,430(31.9) | | 3,464(21.3) | | <0.001 |
| Under college/university degree | 261,435(68.1) | | 12,820(78.7) | |  |
| TDS | -2.33(-3.73, 0.11) | | -1.78(-3.45, 1.19) | | <0.001 |
| Total physical activity (MET-min/day) | 257.7(118.1, 514.2) | | 226.0(94.6, 491.4) | | <0.001 |
| BMI (kg/m2) | 26.5(24.0, 29.5) | | 29.7(26.6, 33.5) | | <0.001 |
| Waist-to-hip ratio | 0.87(0.80, 0.93) | | 0.92(0.86, 0.98) | | <0.001 |
| Random glucose (mg/dL) | 88.2(82.5, 94.3) | | 96.0(87.9, 107.4) | | <0.001 |
| HbA1C (%) | 5.3(5.1, 5.5) | | 6.1(6.0, 6.3) | | <0.001 |
| TC (mg/dL) | 226.5(200.5, 254.4) | | 232.1(203.5, 261.1) | | 0.003 |
| LDL (mg/dL) | 142.2(122.2, 163.8) | | 148.9(127.9, 171.4) | | <0.001 |
| HDL (mg/dL) | 55.7(46.9, 66.3) | | 49.6(42.2, 58.9) | | <0.001 |
| TG (mg/dL) | 126.7(90.2-182.6) | | 170.4(120.0-237.5) | | <0.001 |

Values are presented as median (IQR) for continuous variables or N (%) for categorical variables. Continuous variables were adjusted for age and sex in multiple linear regression model when compare their difference between participants with normal glucose and those with prediabetes. Participants who took lipid-lowering medication were excluded when analyzing TC, TG, LDL and HDL.

Abbreviations: TDS, Townsend Deprivation Score; MET, Metabolic Equivalent of Task; BMI, Body Mass Index; HOMA-B, Homeostatic Model Assessment for Beta Function; HOMA-IR, Homeostatic Model Assessment for Insulin Resistance; OGTT, Oral Glucose Tolerance Test; HbA1C, Hemoglobin A1C; TC, Total Cholesterol; LDL, Low-density Lipoprotein Cholesterol; HDL, High-density Lipoprotein Cholesterol; TG, Triglyceride.

### Supplementary Table 5 Main genetic and phenotypic features of six cluster of prediabetes in HCHS/SOL

| Cluster | Genetic scores | | | | | Metabolic traits | | | | |
| --- | --- | --- | --- | --- | --- | --- | --- | --- | --- | --- |
|  | Beta cell  score | Proinsulin  score | Obesity  score | Lipodystrophy-like score | Liver-lipid score | HOMA-B | HDL-C | BMI | TG | HOMA-IR |
| 1 | Far below Average | Slightly below Average | Above Average | Average | Above Average | Above Average | Slightly below Average | Above Average | Below Average | Above Average |
| 2 | Above Average | Slightly above Average | Below Average | Below Average | Above Average | Far below Average | Above Average | Above Average | Far below Average | Far below Average |
| 3 | Slightly above Average | Average | Far above Average | Below Average | Below Average | Slightly above Average | Slightly below Average | Far above Average | Slightly above Average | Slightly below Average |
| 4 | Slightly above Average | Above Average | Below Average | Above Average | Far below Average | Below Average | Slightly below Average | Far below Average | Far above Average | Below Average |
| 5 | Below Average | Above Average | Below Average | Above Average | Above Average | Slightly above Average | Far below Average | Slightly below Average | Average | Above Average |
| 6 | Slightly above Average | Far below Average | Slightly above Average | Slightly above Average | Above Average | Above Average | Slightly below Average | Below Average | Below Average | Slightly above Average |

### Supplementary Table 6 Baseline characteristics of six clusters of participants with prediabetes in HCHS/SOL

|  | cluster 1 | cluster 2 | cluster 3 | cluster 4 | cluster 5 | cluster 6 | P value |
| --- | --- | --- | --- | --- | --- | --- | --- |
| Number | 488 | 569 | 547 | 742 | 719 | 612 |  |
| Age (year) | 50.0(44.0-58.0) | 51.0(44.0-58.0) | 52.0(46.0-60.0) | 50.0(44.0-57.0) | 50.0(42.0-57.0) | 52.0(46.0-59.0) | 0.618 |
| Sex |  |  |  |  |  |  |  |
| Female | 309(63.3) | 334(58.7) | 333(60.9) | 471(63.5) | 428(59.5) | 356(58.2) | 0.234 |
| Male | 179(36.7) | 235(41.3) | 214(39.1) | 271(36.5) | 291(40.5) | 256(41.8) |  |
| U.S.-born status |  |  |  |  |  |  |  |
| non U.S.-born | 414(84.8) | 484(85.1) | 488(89.2) | 649(87.6) | 618(86.0) | 533(87.2) | 0.237 |
| U.S.-born | 74(15.2) | 85(14.9) | 59(10.8) | 92(12.4) | 101(14.0) | 78(12.8) |  |
| Smoking status |  |  |  |  |  |  |  |
| Never | 295(60.5) | 338(59.4) | 312(57.0) | 466(62.9) | 440(61.3) | 340(55.6) | 0.205 |
| Former | 105(21.5) | 120(21.1) | 123(22.5) | 149(20.1) | 159(22.1) | 136(22.2) |  |
| Current | 88(18.0) | 111(19.5) | 112(20.5) | 126(17.0) | 119(16.6) | 136(22.2) |  |
| Drinking status |  |  |  |  |  |  |  |
| Never | 264(54.1) | 289(50.8) | 281(51.4) | 387(52.2) | 347(48.4) | 320(52.3) | 0.617 |
| Former | 202(41.4) | 255(44.8) | 243(44.4) | 313(42.2) | 327(45.6) | 266(43.5) |  |
| Current | 22(4.5) | 25(4.4) | 23(4.2) | 42(5.7) | 43(6.0) | 26(4.2) |  |
| Hispanic/Latino background |  |  |  |  |  |  |  |
| Central American | 47(9.7) | 54(9.5) | 53(9.7) | 82(11.1) | 87(12.2) | 80(13.1) | <0.001 |
| Cuban | 89(18.3) | 92(16.2) | 131(24.0) | 139(18.8) | 73(10.2) | 168(27.5) |  |
| Dominican | 49(10.1) | 55(9.7) | 63(11.6) | 44(5.9) | 48(6.7) | 66(10.8) |  |
| Mexican | 166(34.2) | 230(40.6) | 164(30.1) | 324(43.8) | 358(50.0) | 151(24.7) |  |
| Puerto Rican | 103(21.2) | 97(17.1) | 106(19.4) | 96(13.0) | 74(10.3) | 107(17.5) |  |
| South American | 32(6.6) | 39(6.9) | 28(5.1) | 55(7.4) | 76(10.6) | 39(6.4) |  |
| Education |  |  |  |  |  |  |  |
| Less Than High School | 197(40.4) | 223(39.2) | 207(37.9) | 278(37.5) | 300(41.8) | 220(36.1) | 0.380 |
| High School or Equivalent | 106(21.7) | 150(26.4) | 126(23.1) | 183(24.7) | 172(24.0) | 150(24.6) |  |
| Greater than High School  or Equivalent | 185(37.9) | 196(34.4) | 213(39.0) | 281(37.9) | 245(34.2) | 240(39.3) |  |
| Annual income |  |  |  |  |  |  |  |
| <$10,000 | 74(16.4) | 85(15.9) | 86(16.9) | 103(15.0) | 84(12.6) | 99(17.6) | 0.336 |
| $10,001-$20,000 | 136(30.2) | 160(29.9) | 161(31.7) | 215(31.2) | 227(34.1) | 172(30.7) |  |
| $20,001-$40,000 | 164(36.4) | 191(35.7) | 169(33.3) | 242(35.2) | 221(33.2) | 184(32.8) |  |
| $40,001-$75,000 | 66(14.6) | 69(12.9) | 64(12.6) | 99(14.4) | 96(14.4) | 86(15.3) |  |
| >$75,000 | 11(2.4) | 30(5.6) | 28(5.5) | 29(4.2) | 37(5.6) | 20(3.6) |  |
| AHEI-2010 | 48.8(43.5, 54.8) | 49.2(44.6-54.5) | 48.7(43.7-54.1) | 49.4(44.0-55.0) | 50.3(45.0-56.2) | 47.7(43.3-52.9) | 0.861 |
| Total physical activity  (MET-min/day) | 171.4(17.1, 828.6) | 205.7(28.6, 720.0) | 171.4(0.0, 600.0) | 162.9(17.1, 685.7) | 194.3(34.3, 840.0) | 108.6(0.0, 610.7) | 0.730 |
| BMI (kg/m2) | 30.1(26.7, 34.7) | 29.9(26.8, 33.3) | 30.2(26.9, 34.2) | 29.1(26.6, 32.6) | 29.6(26.5, 33.5) | 29.5(26.8, 32.6) | <0.001 |
| Waist-to-hip ratio | 0.9(0.9, 1.0) | 0.9(0.9, 1.0) | 0.9(0.9, 1.0) | 0.9(0.9, 1.0) | 0.9(0.9, 1.0) | 0.9(0.9, 1.0) | 0.174 |
| HOMA-Β | 129.8(87.1, 192.7) | 112.5(75.5, 175.8) | 125.7(81.3, 194.0) | 118.9(79.2, 181.5) | 124.2(82.2, 183.7) | 127.7(85.5, 187.9) | 0.257 |
| HOMA-IR | 3.0(1.9, 4.4) | 2.6(1.7, 4.0) | 2.8(1.9, 4.2) | 2.7(1.7, 4.0) | 3.0(1.9, 4.5) | 2.9(1.9, 4.3) | 0.437 |
| Fasting insulin (mU/L) | 12.0(8.5, 18.4) | 10.8(7.3, 16.0) | 11.8(8.0, 17.0) | 11.5(7.0, 16.3) | 12.5(8.0, 18.3) | 12.0(8.0, 18.0) | 0.404 |
| Fasting glucose (mg/dL) | 99.0(92.0, 104.0) | 98.0(92.0, 104.0) | 99.0(92.0, 103.0) | 97.0(92.0, 102.0) | 99.0(92.0, 104.0) | 99.0(93.0, 103.0) | 0.642 |
| OGTT glucose (mg/dL) | 138.0(109.0, 155.0) | 133.0(108.0, 155.0) | 137.0(109.5, 158.0) | 138.0(112.0, 161.0) | 131.0(107.0, 153.2) | 131.0(107.2, 154.8) | 0.395 |
| HbA1C (mmol/mol) | 5.70(5.50, 5.90) | 5.70(5.50, 5.90) | 5.70(5.50, 5.90) | 5.70(5.50, 5.90) | 5.70(5.50, 5.90) | 5.70(5.50, 5.90) | 0.726 |
| TC (mg/dL) | 200.0(174.0, 230.2) | 209.0(179.0, 234.0) | 205.0(179.0, 231.0) | 209.0(184.0, 238.0) | 204.0(180.0, 231.0) | 208.0(180.0, 236.2) | 0.371 |
| LDL (mg/dL) | 126.0(102.0, 150.0) | 129.0(110.0, 155.0) | 127.0(108.0, 150.0) | 131.0(107.0, 155.0) | 128.0(106.0, 148.0) | 131.0(105.2, 156.0) | 0.265 |
| HDL (mg/dL) | 47.0(40.0, 55.0) | 48.0(41.0, 57.0) | 47.0(39.0, 56.0) | 47.0(40.0, 56.0) | 46.0(40.0, 55.0) | 47.0(40.0, 56.0) | 0.277 |
| TG (mg/dL) | 122.0(87.0, 174.2) | 118.0(87.0, 164.0) | 127.0(88.0, 188.5) | 133.5(97.0, 182.8) | 125.0(91.0, 178.0) | 121.0(90.0, 170.2) | 0.299 |
| Health lifestyle score | 5(4, 7) | 6(4,7) | 5(4, 7) | 6(4,7) | 6(4,7) | 5(4, 7) | 0.599 |

Values are presented as median (IQR) for continuous variables or N (%) for categorical variables. ANOVA was used for testing the difference of continuous variables among six prediabetic clusters, and Chi-squared test was used for categorical variable. Participants who took lipid-lowering medication were excluded when analyzing TC, TG, LDL and HDL.

Abbreviations: AHEI-2010, Alternate Healthy Eating Index 2010; MET, Metabolic Equivalent of Task; BMI, Body Mass Index; HOMA-B, Homeostatic Model Assessment for Beta Function; HOMA-IR, Homeostatic Model Assessment for Insulin Resistance; OGTT, Oral Glucose Tolerance Test; HbA1C, Hemoglobin A1C; TC, Total Cholesterol; LDL, Low-density Lipoprotein Cholesterol; HDL, High-density Lipoprotein Cholesterol; TG, Triglyceride.

### Supplementary Table 7 Baseline characteristics of fiver clusters of participants with prediabetes in UKBB

|  | cluster 1 | cluster 2 | cluster 4 | cluster 5 | cluster 6 | P value |
| --- | --- | --- | --- | --- | --- | --- |
| Number | 3,194 | 3,070 | 3,595 | 3,243 | 3,182 |  |
| Age (year) | 62(57-66) | 62(56-66) | 62(57-65) | 62(57-66) | 62(57-66) | 0.640 |
| Sex |  |  |  |  |  |  |
| Female | 1,589(49.7) | 1,513(49.3) | 1,857(51.7) | 1,679(51.8) | 1,571(49.4) | 0.087 |
| Male | 1,605(50.3) | 1,557(50.7) | 1,738(48.3) | 1,564(48.2) | 1,611(50.6) |  |
| Smoking status |  |  |  |  |  |  |
| Never | 1,452(45.7) | 1,472(48.2) | 1,605(44.9) | 1,520(47.0) | 1,473(46.6) | 0.078 |
| Former | 1,189(37.4) | 1,107(36.3) | 1,413(39.5) | 1,215(37.6) | 1,216(38.4) |  |
| Current | 538(16.9) | 474(15.5) | 556(15.6) | 495(15.3) | 475(15.0) |  |
| Drinking status |  |  |  |  |  |  |
| Never | 216(6.8) | 184(6.0) | 197(5.5) | 219(6.8) | 203(6.4) | 0.143 |
| Former | 162(5.1) | 139(4.5) | 195(5.4) | 186(5.7) | 164(5.2) |  |
| Current | 2,813(88.1) | 2,738(89.5) | 3,198(89.1) | 2,832(87.5) | 2811(88.4) |  |
| Education |  |  |  |  |  |  |
| College or University degree | 674(21.1) | 652(21.2) | 793(22.0) | 691(21.3) | 654(20.6) | 0.668 |
| Under college/university degree | 2,520(78.9) | 2,418(78.8) | 2,802(77.9) | 2,552(78.7) | 2,528(79.4) |  |
| TDS | -1.8(-3.5, 1.1) | -1.7(-3.4, 1.3) | -1.9(-3.6, 1.0) | -1.7(-3.4, 1.3) | -1.8(-3.5, 1.1) | 0.760 |
| Total physical activity  (MET-min/day) | 230.6(96.4, 496.3) | 221.3(99.0, 466.9) | 213.4(90.5, 443.9) | 219.0(96.6, 479.1) | 210.9(84.9, 438.0) | 0.082 |
| BMI (kg/m2) | 30.2(26.9, 34.1) | 29.4(26.5, 33.1) | 29.4(26.5, 32.9) | 29.2(26.3, 32.7) | 30.0(26.8, 33.5) | 0.004 |
| Waist-to-hip ratio | 0.92(0.86, 0.98) | 0.92(0.86, 0.98) | 0.92(0.86, 0.98) | 0.92(0.86, 0.97) | 0.92(0.86, 0.98) | 0.423 |
| Random glucose (mg/dL) | 95.4(87.9, 106.2) | 96.1(87.9, 107.2) | 95.1(87.3, 105.8) | 96.0(88.1, 106.1) | 95.7(87.6, 107.5) | 0.859 |
| HbA1C (%) | 6.13(6.05, 6.27) | 6.14(6.05, 6.27) | 6.13(6.05, 6.27) | 6.14(6.05, 6.27) | 6.14(6.05, 6.27) | 0.199 |
| TC (mg/dL) | 208.4(176.5, 243.5) | 210.9(177.9, 244.6) | 211.5(179.5, 247.6) | 206.7(175.4, 243.1) | 207.8(176.3, 242.6) | 0.191 |
| LDL (mg/dL) | 131.1(105.8, 158.2) | 131.3(106.2, 158.5) | 131.3(107.5, 159.5) | 128.6(104.5, 157.2) | 129.9(105.4, 157.8) | 0.238 |
| HDL (mg/dL) | 47.8(40.6, 56.8) | 49.1(41.7, 57.9) | 48.4(41.2, 56.9) | 47.6(40.9, 56.5) | 47.5(40.5, 56.5) | 0.022 |
| TG (mg/dL) | 168.2(118.6, 232.8) | 159.3(110.9, 219.2) | 176.7(125.4, 241.5) | 167.4(121.7, 228.5) | 167.1(118.1, 230.3) | 0.001 |
| Health lifestyle score | 5(3, 6) | 5(4, 6) | 5(4, 6) | 5(4, 6) | 5(3, 6) | 0.236 |

Values are presented as median (IQR) for continuous variables or N (%) for categorical variables. ANOVA was used for testing the difference of continuous variables among five prediabetic clusters, and Chi-squared test was used for categorical variable. Participants who took lipid-lowering medication were excluded when analyzing TC, TG, LDL and HDL.

Abbreviations: TDS, Townsend Deprivation Score; MET, Metabolic Equivalent of Task; BMI, Body Mass Index; HOMA-B, Homeostatic Model Assessment for Beta Function; HOMA-IR, Homeostatic Model Assessment for Insulin Resistance; OGTT, Oral Glucose Tolerance Test; HbA1C, Hemoglobin A1C; TC, Total Cholesterol; LDL, Low-density Lipoprotein Cholesterol; HDL, High-density Lipoprotein Cholesterol; TG, Triglyceride.
